# Study protocol for integrated service delivery to improve adolescent health and HPV vaccination in Lao PDR: A quasi-experimental mixed-methods study

**DOI:** 10.1101/2024.12.08.24318679

**Authors:** Thet Lynn, Vanphanom Sychareun, Kongmany Chaleunvong, Viengnakhone Vongxay, Suksamone Thongmyxay, Chandavieng Phommavong, Chansathith Taikeophithoun, Sengtavanh Keokenchanh, Josien Van Der Kooij, Bangyuan Wang

**Affiliations:** Asia Programme, Health Poverty Action (HPA), Vientiane Capital, Lao PDR; Faculty of Public Health, University of Health Science (UHS), Vientiane Capital, Lao PDR

## Abstract

1.

**Background:** The Lao People’s Democratic Republic (PDR) faces the challenge of high cervical cancer incidence and low HPV vaccine coverage, alongside broader adolescent health concerns like high adolescent pregnancy rates and inadequate Sexual and Reproductive Health (SRH) literacy. The integration of HPV vaccination with adolescent health services aims to address these issues comprehensively, increasing vaccine uptake and improving overall adolescent health outcomes.

**Methods:** This study will employ a quasi-experimental design with mixed methods to assess the impact of integrating HPV vaccination with adolescent health services in two districts of Louangnamtha province in Laos. The study population includes girls aged 10-13 years for HPV vaccination and boys and girls aged 10-13 years for other adolescent health services. The intervention district will receive an integrated service package comprising HPV vaccination and adolescent health services delivered through school-based, facility-based, and outreach approaches. The comparison district will continue with its standard HPV vaccination and routine adolescent health service delivery. The intervention will be for 7 months from 2024 November to 2025 May. Quantitative data on HPV vaccine uptake, SRH knowledge and attitudes, and utilization of adolescent health services will be collected at baseline, midline, and endline. Qualitative data on barriers and facilitators to implementation, provider workload and capacity, and stakeholder perceptions will be collected through in-depth interviews and focus group discussions at endline.

**Results:** The study outcome is the evidence on the impact of integration on change in HPV vaccine uptake, change in SRHR knowledge and attitude, utilization of adolescent health services, healthcare providers’ workload and capacity, and stakeholders’ opinions regarding the integrated intervention.

**Recommendations:** The study’s findings will inform policies and practices in Lao PDR.

**Study Registrations:**

1) ClinicalTrials.gov ID NCT06650956,
2) OSF Pre-registration: https://doi.org/10.17605/OSF.IO/MGEQ7

## 2. Introduction

Lao People’s Democratic Republic has a population of around 7.6 million in 2023. It faces a significant health challenge with neoplasms being the third leading cause of death. Among females aged 15-49, cervical cancer is the second leading cause of death due to neoplasms in 2021.(1,2) This trend has persisted since 2010, underscoring the need for sustainable and effective preventative measures. (3,4) The introduction of prophylactic human papillomavirus (HPV) vaccines offers a promising solution for the primary prevention of cervical cancer and other HPV-related diseases. However, the global impact of these vaccines, particularly in low- and middle-income countries (LMICs) like Laos, hinges on successful implementation strategies. These strategies must encompass policy development, financing, effective communication, public health program delivery, and widespread acceptance. Despite the availability of HPV vaccines and the establishment of national vaccination programs in many countries across the world, equitable access and optimal utilization remain critical challenges, especially in LMICs where cervical cancer screening programs are often limited or non-existent.(5)

To address this global health challenge, the World Health Organization (WHO) has outlined a global strategy to eliminate cervical cancer as a public health problem. This strategy prioritizes the vaccination of adolescent girls against HPV, aiming to achieve 90% HPV vaccination coverage among girls by age 15. However, the inequitable distribution of HPV vaccine coverage, with higher rates in higher-income countries compared to LMICs, remains a significant concern. To bridge this gap, the WHO advocates for a multifaceted approach, including securing sufficient and affordable HPV vaccines, increasing vaccination coverage and quality, improving communication and social mobilization efforts, and fostering innovation in vaccine delivery.(6)

Despite Lao PDR’s significant economic growth and improvements in population health over the past two decades, the issue of health equity is particularly salient among adolescents.(7) One of the most pressing public health concerns in Lao PDR is the high rate of adolescent pregnancy, the highest in Southeast Asia.(8) The COVID-19 pandemic has further exacerbated existing health inequities in Lao PDR. The pandemic’s economic impact has pushed more households into poverty, disproportionately affecting vulnerable populations, including adolescents. Such issues are beyond health concerns and are social ones, often leading to school dropout, limited prospects for young mothers, and adverse health outcomes for both mothers and infants.(9) Early marriage, limited access to sexual and reproductive health (SRH) information and services, and inadequate comprehensive sexuality education (CSE) all contribute to this problem in Lao PDR. A study conducted in 2017 revealed that 65.5% of school-going adolescents in Lao PDR had inadequate SRH literacy, further highlighting the need for improved access to information and services. (10,11) In addition, the utilization of government-managed health centres, which serve as the entry point for primary care, is low due to perceptions of lower quality care compared to hospitals and private clinics.(12) HPV vaccination is a safe and effective way to prevent cervical cancer. Yet vaccine coverage in Lao PDR remains suboptimal. In Laos, HPV vaccination programme was first introduced in 2020 to girls aged 10-17, whether or not they are enrolled in school. Cumulatively from 2020 to 2023, nearly 60% of 10-17-year-old-girls have completed 2 doses of HPV vaccine. Starting from 2024, Laos will switch to a single-dose HPV vaccination strategy targeting at 10-13-year-old adolescent girls.(13)

### Rationale for Integration

The integration of HPV vaccination with adolescent health services is a strategic approach that aligns with the Lao PDR’s national health priorities and international recommendations. This integration is not merely a matter of convenience but a necessity to address the complex and interconnected health needs of adolescents.

Firstly, integrating HPV vaccination with adolescent health services can significantly increase vaccine uptake. By offering the vaccine alongside other essential health services, such as SRHR education, counselling, and other age-appropriate youth friendly services, healthcare providers can reach a larger number of adolescents, including some who may not specifically seek out HPV vaccination. This approach is particularly important in Lao PDR, where access to healthcare services can be limited, especially in rural areas.

Secondly, the integrated approach can improve adolescent health outcomes more broadly. By providing comprehensive SRH services, including HPV vaccination, adolescents can receive the necessary information and care to make informed decisions about their sexual and reproductive health. This can lead to a reduction in unintended pregnancies, STIs, and other adverse health outcomes.

Thirdly, integrating HPV vaccination with adolescent health services can help address the social and cultural factors that contribute to poor health outcomes among adolescents. By creating a safe and supportive environment where adolescents can access a range of health services, healthcare providers can build trust and rapport with this vulnerable population. This can facilitate open communication about sensitive topics, such as sexuality and gender-based violence, and empower adolescents to take control of their health.

Finally, the integrated approach can be a cost-effective way to deliver essential health services to adolescents. By leveraging existing healthcare infrastructure and personnel, the government can maximize the impact of its investments in adolescent health. Additionally, the prevention of cervical cancer through HPV vaccination can lead to significant cost savings in the long run by reducing the need for expensive cancer treatment.

### Social Ecological Model Theory

The design of the integration is influenced by the Social Ecological Model Theory. This model recognizes the interconnectedness of individual, interpersonal, organizational, community, and policy-level factors in shaping health behaviours and outcomes. *At the individual level*, the integration aims to increase HPV vaccine uptake by making it more accessible and convenient for adolescent girls. By offering the vaccine alongside other essential health services, the initiative acknowledges that adolescents’ health needs extend beyond HPV prevention and seeks to address them comprehensively. *On an interpersonal level*, the integration recognizes the influence of family, peers, and healthcare providers on adolescents’ health decisions. By involving caregivers and providing education and counselling, the initiative aims to create a supportive environment that encourages vaccine uptake and promotes positive health behaviours. *At the organizational level*, the integration leverages existing education and healthcare infrastructure and personnel to maximize the reach and efficiency of service delivery. This approach acknowledges the importance of healthcare systems in facilitating access to and utilization of health services. *Within the community*, the integration aims to address social and cultural factors that may influence vaccine hesitancy or hinder access to care. By engaging community leaders and tailoring interventions to local contexts, the initiative seeks to build trust and promote acceptance of HPV vaccination. Ultimately, *at the policy level*, the integration aligns with national health priorities and international recommendations for HPV prevention and adolescent health. By embedding HPV vaccination within a broader framework of adolescent health services, the initiative contributes to the long-term goal of improving health equity and reducing the burden of cervical cancer in Lao PDR.

Integrating HPV vaccination with adolescent health services presents a unique opportunity to address these multifaceted challenges.(14) By leveraging existing healthcare platforms and reaching adolescents during a critical period of their development, this integrated approach can potentially increase HPV vaccine uptake, improve adolescent health outcomes, and contribute to the broader goal of achieving health equity in Laos.

## 3. Materials and Methods

### 3.1 Objectives

The overall objective of the study is to determine how integrating HPV vaccination services with adolescent health services could increase HPV vaccine coverage equitably and sustainability, as well as contribute to selected adolescent health outcomes in Lao.

#### Specific Objectives (SO)

1. To determine the extent to which the integration of HPV vaccination with adolescent health services increases HPV vaccine uptake among 10-13-year-old girls in the intervention district compared to the comparison district over 7 months.
2. To assess the impact of the integrated intervention on adolescents’ knowledge, attitudes, and practices related to sexual and reproductive health (SRH) among 10-13-year-old boys and girls, as well as their utilization of other adolescent health services.
3. To identify barriers and facilitators to implementing the integrated intervention in the Laotian context, including its impact on healthcare providers’ workload and capacity.
4. To explore the perceptions of adolescents, caregivers, healthcare providers, and other stakeholders regarding integrated intervention, including its perceived benefits and challenges.
5. To determine the costs and effects of integrating HPV vaccination with adolescent health services compared to delivering HPV vaccination alone and assess the cost-effectiveness of the integrated intervention in the Laotian context.

#### Research Questions

##### SO-1

1.1 What is the difference-in-differences (DID) of change in the HPV vaccine uptake in the 10-13-year-old girls’ population between the intervention and comparison districts over 7 months?
1.2 What factors contribute to the differences – if any – in HPV vaccine uptake as such between the intervention and comparison districts?

##### SO-2

2.1 How does the integrated intervention affect 10-13-year-old adolescents’ knowledge, and attitudes related to SRHR in the intervention district compared to the comparison district over 7 months?
2.2 What changes in the utilization of adolescent health services among 10-13-year-olds are observed in the intervention district compared to the comparison district?

##### SO-3

3.1 What are the main barriers and facilitators to implementing the integrated intervention in the Lao context?
3.2 How does the integrated intervention impact healthcare providers’ workload and capacity in the intervention district?

##### SO-4

4.1 What are the perceptions of adolescents, caregivers, healthcare providers, educators and other key stakeholders regarding the benefits and challenges of the integrated intervention?
4.2 How do the perceived benefits and challenges of the integrated intervention vary among different stakeholder groups?

##### SO-5

5.1 What are the costs and effects of integrating HPV vaccination with adolescent health services compared to delivering HPV vaccination alone?
5.2 How does the cost-effectiveness of the integrated intervention compare to that of delivering HPV vaccination alone in the Lao context?

#### Study Design

This study will employ a quasi-experimental design with mixed methods to assess the impact of integrating HPV vaccination with adolescent health services in Lao PDR. A quasi-experimental design is appropriate as random assignment of participants to intervention and comparison groups is not feasible in this implementation setting. The intervention model is a parallel assignment, where one group receives the intervention, i.e., integrated HPV and SRH education services, and the other group does not, i.e., standard HPV vaccine service without any integration. Masking is used in this study to prevent bias. Only the outcomes assessor is masked, meaning they do not know which group received the intervention. The mixed methods approach will allow for an in-depth understanding of the quantitative impact of the intervention on HPV vaccine uptake and other health outcomes, as well as the qualitative aspects of feasibility, acceptability, and barriers/facilitators to implementation.

#### Study Settings

The study will be conducted in two districts of Louangnamtha province in Lao PDR: one intervention district and one comparison district. A lower HPV vaccine coverage of province, availability of adolescent health services, and willingness of local authorities and healthcare providers to participate influences the selection of intervention district. The comparison district will be selected based on similar characteristics to ensure comparability. Comparison district will continue with its standard HPV vaccination (targeting 10-year-old girls) and routine adolescent health service delivery, which may not include the same level of integration or the full range of SRHR services offered in the intervention district.

As shown in figure-2, HPV1 vaccination coverage among 10-17 year old girls in Laos in 2023 varies by province, with Bolikhamxai leading at 27.6% and Bokeo trailing at 12%. The national average stands at 17.3%. While Louangnamtha’s 12.3% coverage exceeds the lowest rates, it remains below the national average, suggesting a need for targeted efforts to improve vaccine uptake in this region and reach the goal of preventing cervical cancer.(13)

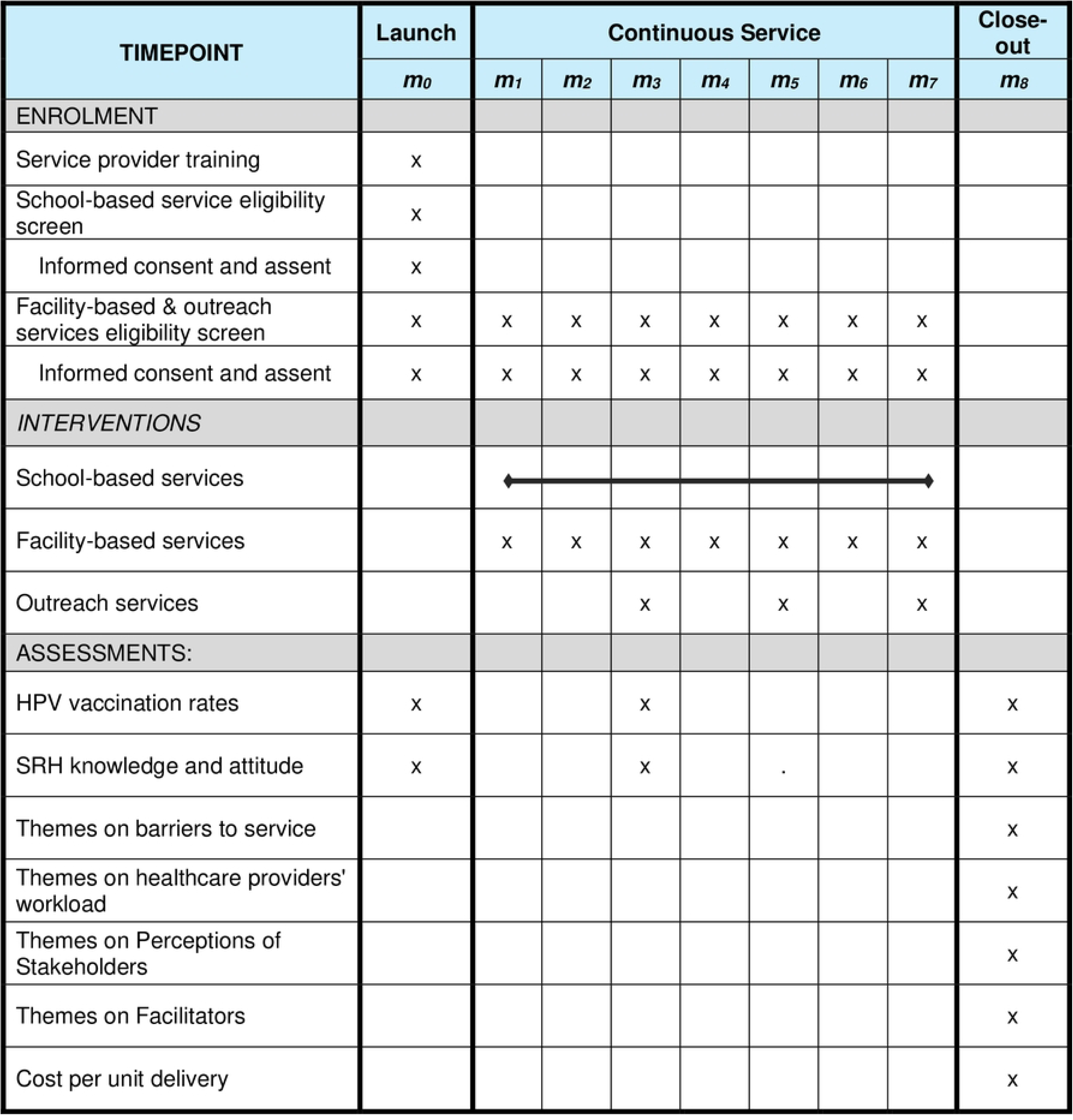

**Figure 2:**
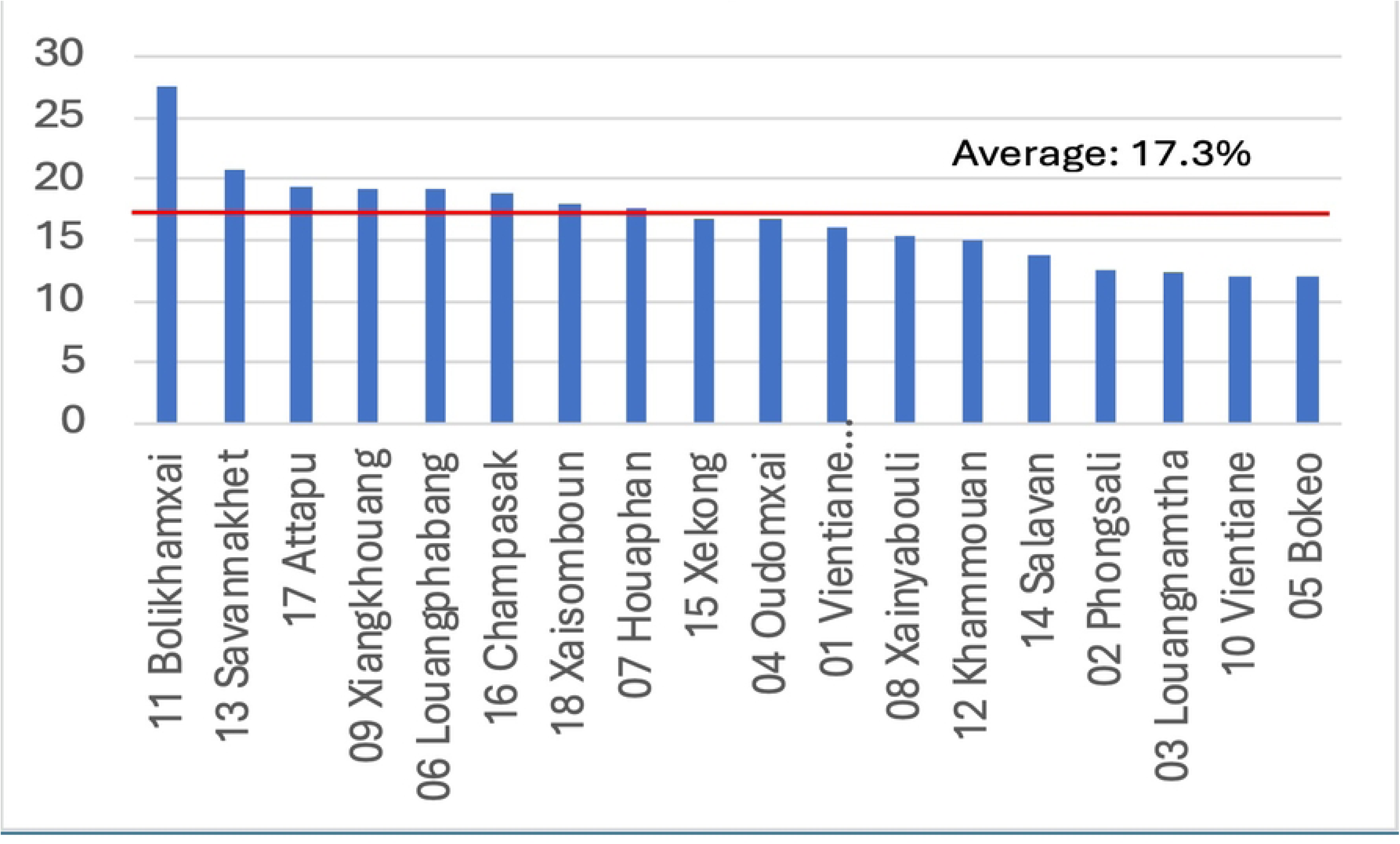
HPV1 coverage by province, Laos, 2023

Louangnamtha province has achieved good progress in reproductive health, with high rates of modern contraceptive use and demand for family planning satisfied. This suggests a positive environment for integrating HPV vaccination with existing reproductive health services. Child nutrition indicators have also improved, particularly in reducing underweight prevalence, indicating a potential opportunity to leverage child health services for HPV vaccine delivery. However, immunization coverage has slightly declined, and disparities exist in hepatitis B birth dose coverage compared to BCG, highlighting the need for targeted interventions to improve vaccine uptake, especially among specific populations and geographic areas.(15)

The study will be conducted in two districts within Louangnamtha province: Sing (intervention) and Viengphoukha (comparison). These districts were purposively selected based on low coverage of HPV vaccination:

- Investigator convenience: The research team’s familiarity with these districts facilitates logistical arrangements and coordination.
- Lower HPV vaccine coverage: Both districts exhibit relatively lower HPV vaccination rates, aligning with the study’s aim to assess the impact of the integrated intervention in areas with greater need for improvement.
- SRHR program opportunities: The districts present promising opportunities for integrating HPV vaccination with existing sexual and reproductive health (SRH) programs, enhancing the potential for successful implementation and impact.

### 3.2 Study Population

#### 3.2.1 Service users

- *HPV Vaccination*: The primary target population for the HPV vaccine will be 10-year-old girls residing in the selected districts. The age range of 10-13 years is set to allow operational flexibility in a context where vital registries are weak and missed opportunities for vaccination are prevalent as indicated in anecdotal reports. This population group will be studied to determine the extent to which the integration of HPV vaccination with adolescent health services increases HPV vaccine uptake among those in the intervention district compared to the comparison district over 7 months.
- *Adolescent Health Service:* The target population for the other adolescent health services will include both boys and girls aged 10-13 years residing in the selected districts. This population will be studied to measure the effect of the integrated intervention on adolescents’ knowledge and attitudes related to their sexual and reproductive health (SRH). They will be the key informants for the study to explore their perceptions regarding the integrated intervention, including its perceived benefits and challenges. In-depth interviews and focus group discussions will be conducted to capture these insights.
- Inclusion criteria The inclusion criteria for the study are as follows:

- Females aged 10-13 years who are eligible for HPV vaccination and will receive sexual and reproductive health (SRH) education.
- Males aged 10-13 years who will receive SRH education.
- Male and female students aged 10-13 years enrolled in randomly selected schools with at least 100 students in the target classes (year 5 primary and year 1, 2, and 3 secondary).
- Out-of-school males and females aged 10-13 years living in two remote villages within the catchment area of each of four randomly selected health centers located near the selected schools.
- Exclusion criteria The exclusion criterion for the study is as follows:

- Males and females younger than 10 years or older than 13 years.

#### 3.2.2 Service providers

*Front-line healthcare providers*: Front-line Healthcare providers including primary and community health cadre who are providing the integrated service package will be important informants to the study for identification of barriers and facilitators to implementing the integrated intervention, including its impact on healthcare providers’ workload and capacity. Interviews and focus group discussions will be conducted to gather qualitative data on these aspects.
*School teachers*: Primary school teachers teamed up with front-line healthcare providers in providing comprehensive sexuality education (CSE) will play pivotal role in contributing their observations and opinions towards learning how the intervention is designed, implemented as well as the improvement areas.

#### 3.2.3 Stakeholders

- *Caregivers or guardians of adolescents* will be invited to participate in the study to share their perceptions regarding the integrated intervention, including its perceived benefits and challenges. In-depth interviews and focus group discussions will be conducted to capture these insights.
- *Community leaders*: It is important to understand their views on the integrated intervention and its impact on the community. Their perceptions will be gathered through qualitative methods to identify common themes and differences across stakeholder groups.
- *Policymakers and program managers* will be involved in interviews and focus group discussions to provide insights into the feasibility, acceptability, and potential policy implications of the integrated intervention.

### 3.3 Sample and Sampling

#### 3.3.1 Sample and sampling matrix – Quantitative and qualitative approaches

**Table 1.**
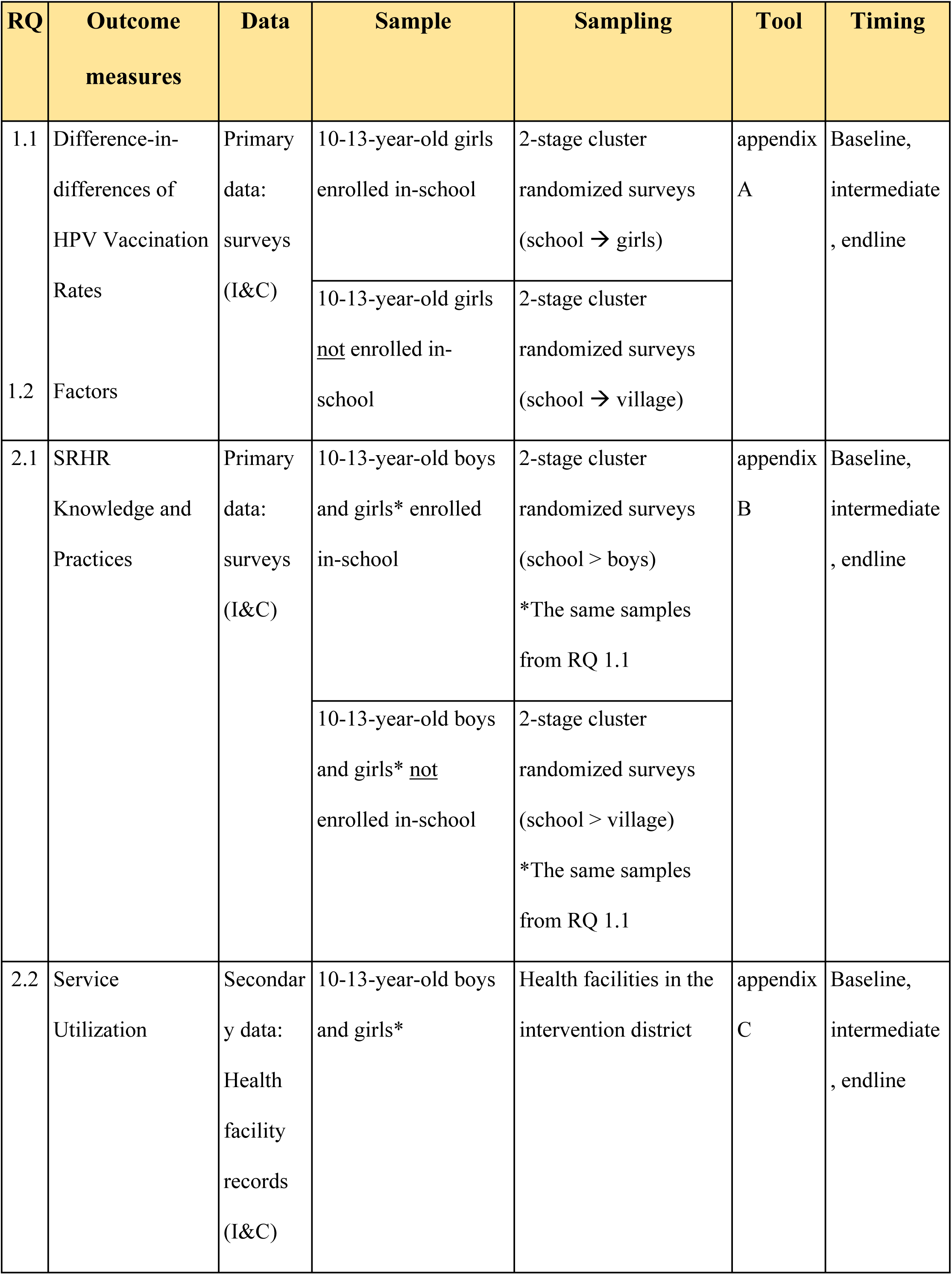
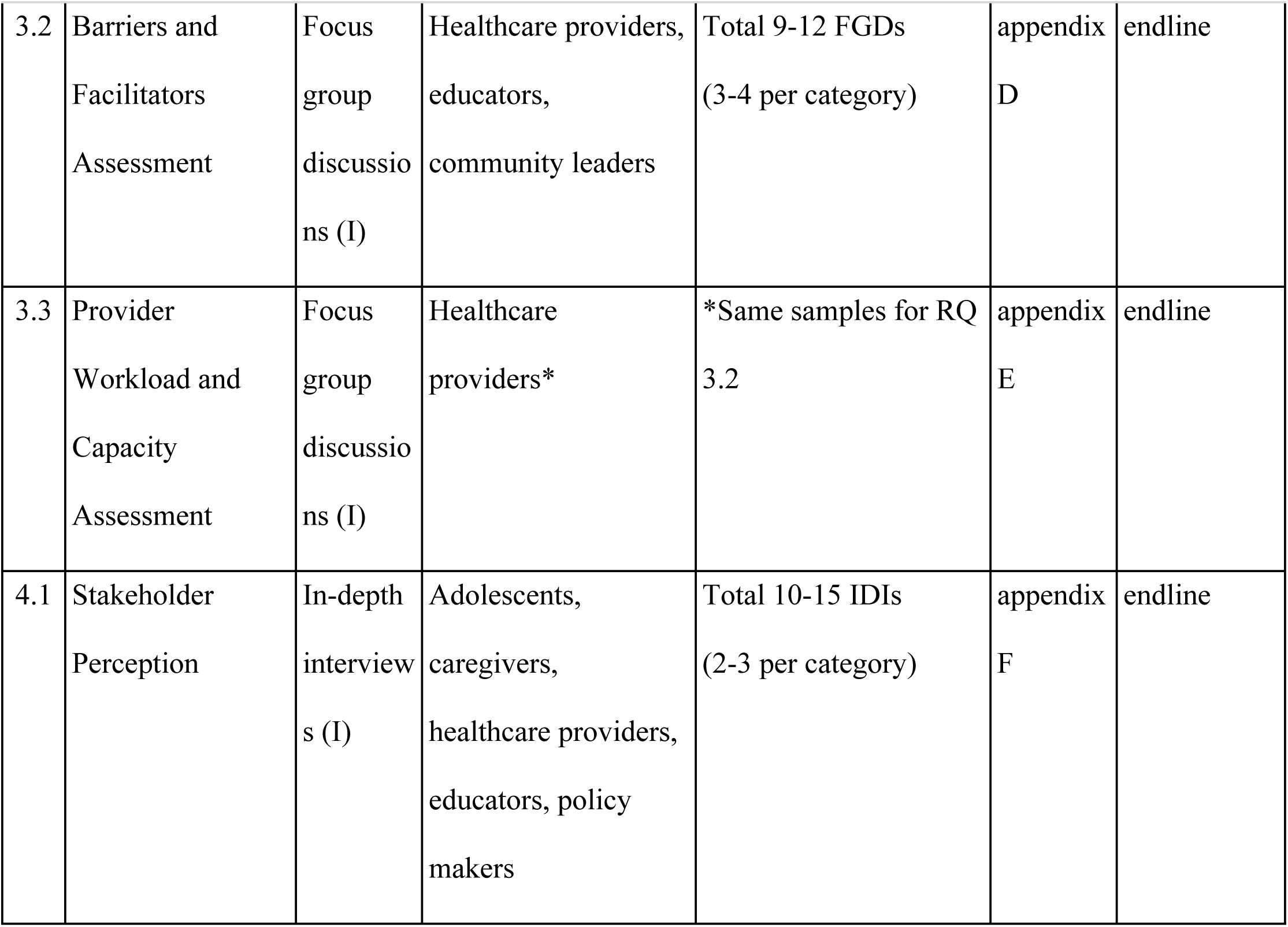
Sample and sampling matrix. I = Intervention district; C = Comparison district

| RQ | Outcome measures | Data | Sample | Sampling | Tool | Timing |
| --- | --- | --- | --- | --- | --- | --- |
| 1.1 | Difference-in-differences of HPV Vaccination Rates | Primary data: surveys (I&C) | 10-13-year-old girls enrolled in-school | 2-stage cluster randomized surveys (school → girls) | appendix A | Baseline, intermediate, endline |
| 1.2 | Factors |  | 10-13-year-old girls <u>not</u> enrolled in-school | 2-stage cluster randomized surveys (school → village) |  |  |
| 2.1 | SRHR Knowledge and Practices | Primary data: surveys (I&C) | 10-13-year-old boys and girls* enrolled in-school | 2-stage cluster randomized surveys (school > boys)<br>*The same samples from RQ 1.1 | appendix B | Baseline, intermediate, endline |
|  |  |  | 10-13-year-old boys and girls* <u>not</u> enrolled in-school | 2-stage cluster randomized surveys (school > village)<br>*The same samples from RQ 1.1 |  |  |
| 2.2 | Service Utilization | Secondary data: Health facility records (I&C) | 10-13-year-old boys and girls* | Health facilities in the intervention district | appendix C | Baseline, intermediate, endline |
| 3.2 | Barriers and Facilitators Assessment | Focus group discussions (I) | Healthcare providers, educators, community leaders | Total 9-12 FGDs (3-4 per category) | appendix D | endline |
| 3.3 | Provider Workload and Capacity Assessment | Focus group discussions (I) | Healthcare providers* | *Same samples for RQ 3.2 | appendix E | endline |
| 4.1 | Stakeholder Perception | In-depth interviews (I) | Adolescents, caregivers, healthcare providers, educators, policy makers | Total 10-15 IDIs (2-3 per category) | appendix F | endline |

#### 3.3.2 Sample size for quantitative approach

The study will involve young adolescent girls and boys aged 10-13 years old, both in-school and out-of-school. The sample size will be determined using the formula for proportional studies in a quasi-experimental design, with a design effect of 2 to account for sampling methods other than simple random sampling. The sample size for boys will be equal to that of girls.

The formula used is as follows:

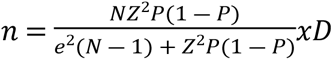

Where: n = number of required samples,
N = known number of study population,
Z = level of confidence at 95% (with a value of 1.96 for alpha 0.05),
e = precision of event measurement (or margin of error),
P = prevalence or proportion of event,
D = design effect

The prevalence of HPV1 vaccination coverage in Louangnamtha province (P = 12.3%) and the total number of girls aged 10-13 years old (N = 3,364) will be used for the calculation.

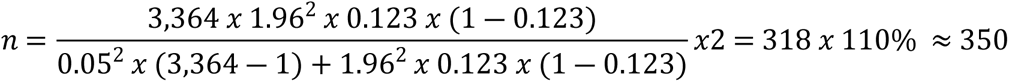

The estimated sample size for girls is 350, including a 10% buffer. The sample size for boys will be equal to that of girls (350). The total sample size for both intervention and comparison sites will be 700 girls and boys aged 10-13 years old.

#### 3.3.3 Sampling for quantitative approach

The study will involve two districts (Sing and Viengphoukha) in Luangnamtha province, pre-assigned as intervention and comparison districts. The sampling strategy will target year 5 primary school and year 1 secondary school students, with 40% of the sample (140 students: 70 girls, 70 boys) from year 5 and 50% (175 students: 88 girls, 87 boys) from year 1. The remaining 10% (35 students: 18 girls, 17 boys) will be out-of-school adolescents, potentially missed by school vaccination programs.

The sampling strategy will involve the following steps:

- *School Selection*: In each district, primary and secondary schools with at least 100 students in the target classes (year 5 primary, year 1, 2 and 3 secondary) will be identified. From this list, four primary schools and four secondary schools will be randomly selected to participate in the study.
- *School Adolescents Recruitment*: Within each selected school, two lists will be created: one for girls and one for boys aged 10-13 years. Each student on these lists will be assigned an ID number. A sex-stratified systematic sampling approach, proportional to the size of each list, will be employed to select participants. The sampling interval will be calculated as the total number of students on the list divided by the desired sample size. The first participant will be randomly selected, and subsequent participants will be chosen based on the sampling interval until the desired sample size is reached. Informed consent will be obtained from selected students and their parents/guardians before conducting structured interviews.
- *Out-of-School Adolescents Recruitment*: Out-of-school adolescents, who may have missed school-based vaccination opportunities, will be recruited through purposive sampling. The process will begin by randomly selecting four health centres (HCs) located near the eligible schools in each district. Within each HC’s catchment area, two villages from zones 2 and 3 (mobile and outreach zones) will be selected based on the highest number of girls missing vaccination or out of school. Village health volunteers and HC chiefs will assist in identifying and recruiting eligible girls and boys within these villages. All identified out-of-school adolescents within the selected villages will be recruited for the study.

#### 3.3.4 Sample size and sampling for qualitative approach

The qualitative sampling in this study will employ purposive and convenience sampling methods to recruit participants who possess the necessary knowledge and experience to provide valuable insights. The recruitment process will involve the following:

- *Healthcare providers*: The research team will collaborate with healthcare facilities in the intervention district to identify and recruit healthcare providers who have been actively involved in delivering the integrated service package. The selection will prioritize those with diverse roles and experiences in providing adolescent health services and HPV vaccination.
- *Educators*: The research team will coordinate with schools in the intervention district to identify and recruit teachers who have participated in the CSE training and have experience delivering CSE sessions to students. The selection will focus on teachers who have actively engaged with the integrated intervention and can provide insights into its implementation and impact.
- *Community leaders*: The research team will engage with local authorities and community organizations to identify and recruit influential community leaders who involve in women health promotion. The selection will prioritize leaders who can provide insights into community dynamics, cultural factors, and potential barriers or facilitators to the intervention’s success.
- *Adolescents*: The research team will collaborate with schools and community organizations in the intervention district to identify and recruit adolescent boys and girls aged 10-13 years who have participated in the integrated intervention. The selection will aim for diversity in terms of gender, socioeconomic background, and experiences with the intervention.
- *Caregivers*: The research team will utilize existing networks and community contacts to identify and recruit caregivers of adolescents who have participated in the integrated intervention. The selection will prioritize caregivers whose daughter missed the previous HPV vaccination, or whose daughter was vaccinated at age 12 or above.
- *Policymakers:* The research team will leverage their existing relationships with government officials and policymakers to identify and recruit individuals who are involved in decision-making related to adolescent health and vaccination programs. The selection will prioritize those who can provide insights into the policy implications of the integrated intervention and its potential for scalability and sustainability, or who directly involve in vaccination program/planning.

The recruitment process will involve providing potential participants with information about the study’s purpose, procedures, and potential benefits and risks. Informed consent will be obtained from all participants or their legal guardians before interview and before audio recording. The research team will ensure that participants understand their rights, including the right to withdraw from the study at any time without penalty.

The qualitative data collection will involve conducting an estimated 10-15 focus group discussions (FGDs) to explore the barriers and facilitators encountered during the intervention, as well as its impact on the workload and capacity of healthcare providers. Additionally, 20-25 in-depth interviews (IDIs) will be carried out with various stakeholders, including caregivers of adolescents, healthcare and education providers, and policymakers. The principle of information saturation will guide the determination of the final number of interviews and discussions. If saturation is reached before the estimated numbers, data collection/interviews may cease earlier. Conversely, if additional insights are needed, further interviews or discussions may be conducted beyond the initial estimates.

### 3.4 Study Procedures

To answer the research questions, a mixed-methods approach will be employed, utilizing quantitative followed by qualitative data collection and analysis techniques. *Table 1. Sample and sampling matrix*

#### 3.4.1 Quantitative Data

- *HPV Vaccine Uptake*: To assess the impact of the integrated intervention on HPV vaccine uptake (research question 1.1), vaccination coverage rates among 10-13-year-old girls in both the intervention and comparison districts will be measured at baseline, at 3^rd^ month and end of intervention. Primary data from the study population of adolescent girls’ cohort of 10-13-year-old age. Administrative data from immunization registries and health facility records will be used to triangulate the vaccination coverage.
- *Knowledge and Attitudes (KA) Surveys*: To evaluate the impact of the integrated intervention on adolescents’ knowledge and attitudes related to SRHR (research question 2.1), KA surveys will be administered to a representative sample of 10-13-year-old boys and girls in both the intervention and comparison districts at baseline and endline.
- *Utilization of Adolescent Health Services*: To examine changes in the utilization of other adolescent health services (research question 2.2), data on service utilization (e.g., number of visits for counselling, education) will be collected from health facilities in both districts at baseline and endline.

#### 3.4.2 Qualitative Data

- *Barriers and Facilitators*: To identify barriers and facilitators to implementation (research question 3.2), in-depth interviews and focus group discussions will be conducted with healthcare providers, educators, community leaders, and other key stakeholders involved in the intervention.
- *Provider Workload and Capacity:*: To assess the impact of the integrated intervention on healthcare providers’ workload and capacity (research question 3.3), interviews and focus group discussions will be conducted with healthcare providers in the intervention district.
- *Perceptions of Stakeholders*: To explore the perceptions of adolescents, caregivers, healthcare providers, educators, and other stakeholders regarding the integrated intervention (research questions 4.1 and 4.2), in-depth interviews and focus group discussions will be conducted with representatives from each group.

### 3.5 Intervention

The intervention is an integrated service package comprised of HPV vaccination and adolescent health services in the intervention district, delivered through multiple touchpoints. The design of the integrated approach is informed by evidence from successful interventions in other settings, such as Togo’s experience integrating health education with HPV vaccination. (14)

The integrated service package will be delivered through three primary touchpoints – school-based, health facility based and outreach, drawing on evidence from various studies that have shown the effectiveness of multi-component interventions. (16)

#### 3.5.1 School-Based Service Delivery

- *HPV Vaccination*: Girls aged 10-13 years will receive the HPV vaccine at school in the last week of 2024 November. A vaccination campaign will be conducted in primary schools to promote HPV vaccination and other relevant vaccinations for girls. The campaign will use HPV vaccine promotion materials from Lao Ministry of Health.
- *Comprehensive Sexuality Education (CSE)*: A refresher training will be provided to service providers of CSE, i.e, teachers in schools. Teacher’s training manual on CSE will be used, covering topics such as puberty, menstrual hygiene, STIs, HPV, vaccination, cervical cancer, healthy relationships, risk behaviors and self-protection. Two-to-three-day long training will take place in 2024 September. Materials for references will be the International Technical Guidance on Sexuality Education (UNESCO 2018), Girl’s Project training manual, Vientiane Youth Center (VYC) outreach manual, presentation slides and posters about HPV infection and vaccine.

CSE sessions will be delivered to primary school students aged 10-13 years. The sessions will cover a range of SRHR topics and will be conducted once every three weeks for a total of 10-12 sessions in the school year from 2024 November to 2025 May. CSE sessions will cover a broad range of SRHR topics including such as puberty, menstrual hygiene, STIs, HPV, vaccination, cervical cancer, healthy relationships, risk behaviors and self-protection. Interactive activities, using existing and appropriated photos, short-videos and student-based-approach will be used to conduct CSE session by trained teacher in target primary schools. The sessions will be aided by relevant audio-visual learning contents.

- *Follow-Up*: The site project coordinator will join CSE sessions in schools monthly to monitor progress.

#### 3.5.2 Facility-Based Service Delivery

- *HPV Vaccination*: Girls aged 10-13 years will receive the HPV vaccine at health facilities during regular clinic hours. This will occur throughout the months from 2024 November to 2025 May.
- *SRHR Services*: Healthcare providers will receive refresher training on HPV vaccination, SRHR-relevant services, adolescent and youth-friendly services (AYFS), and counseling skills. The training will utilize the National Adolescent and Youth Friendly Service Guidelines, Vientiane Youth Center (VYC) outreach manual, and posters about HPV infection and vaccine. Health facilities will provide a full range of SRHR services, including education, counseling, contraception, and disease prevention and management. They will utilize the National Adolescent and Youth Friendly Service Guidelines, posters about HPV infection and Vaccine, worksheets, record sheets, and report formats.
- *Follow-Up*: The site project coordinator will visit health facilities monthly to monitor the provision of service in the intervention package.

#### 3.5.3 Community Outreach

- *HPV Vaccination*: Girls aged 10-13 years will receive the HPV vaccine through community outreach campaigns. This will occur quarterly from 2024 November to 2025 May.
- *SRHR Promotion and Service Delivery*: SRHR promotion messages and HPV vaccines will be delivered to out-of-school girls through existing community outreach activities. This will occur quarterly from the third quarter of 2024 to the second quarter of 2025. The outreach will utilize the Vientiane Youth Center (VYC) outreach manual, HPV infection and vaccine messages and posters.
- *Follow-Up*: The site project coordinator will visit during outreach sessions quarterly to monitor progress.

#### 3.5.4 Standard HPV Vaccination (Comparison District)

- In the comparison district, routine HPV vaccination services will be provided to girls aged 10-13. A school-based vaccination campaign will be conducted in 2024 November to promote and administer the HPV vaccine. Additionally, the vaccine will be available at health facilities starting in 2024 November. Outreach activities will also be conducted to ensure access to the vaccine for hard-to-reach populations and out-of-school girls.
- Unlike the intervention district, there will be no additional training for nor service by healthcare providers on comprehensive sexuality education (CSE) or other sexual and reproductive health (SRH) services, and there will be no dedicated community outreach sessions on SRH topics.

### 3.6 Participant timeline

The study is expected to begin participant recruitment in November 2024 and complete recruitment by May 2025. Data collection is estimated to start in November 2024, with the primary completion date, i.e., final data collection date for the primary outcome measure, expected in May 2025. The study completion date is estimated to be in May 2025. See figure-1: SPIRIT schedule of study stages of enrolment, intervention, outcome assessment.

### 3.7 Outcome Measures

The study will assess the outcome measures as mentioned in detail in table 2 – outcome measures.

**Table 2.**
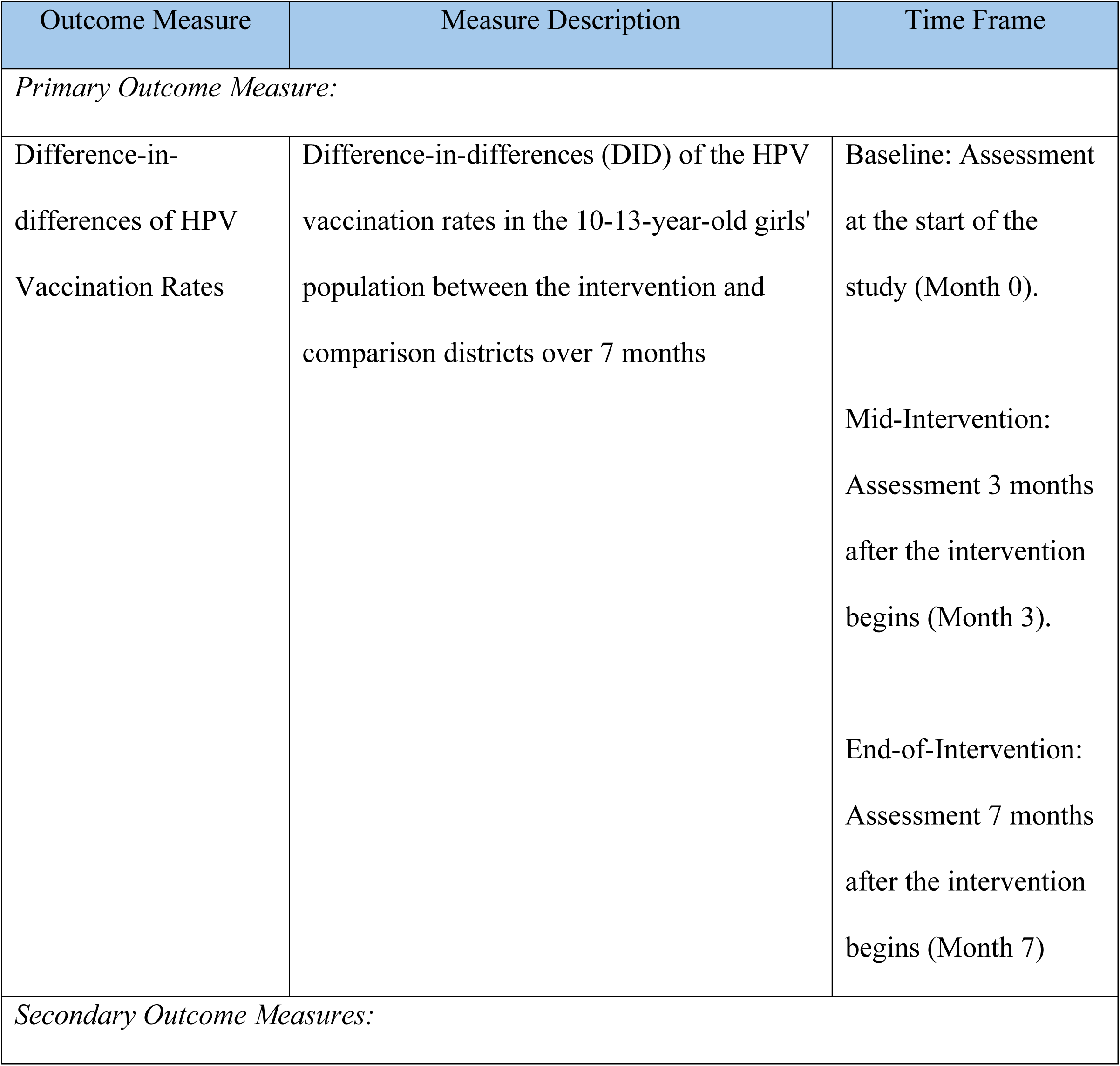
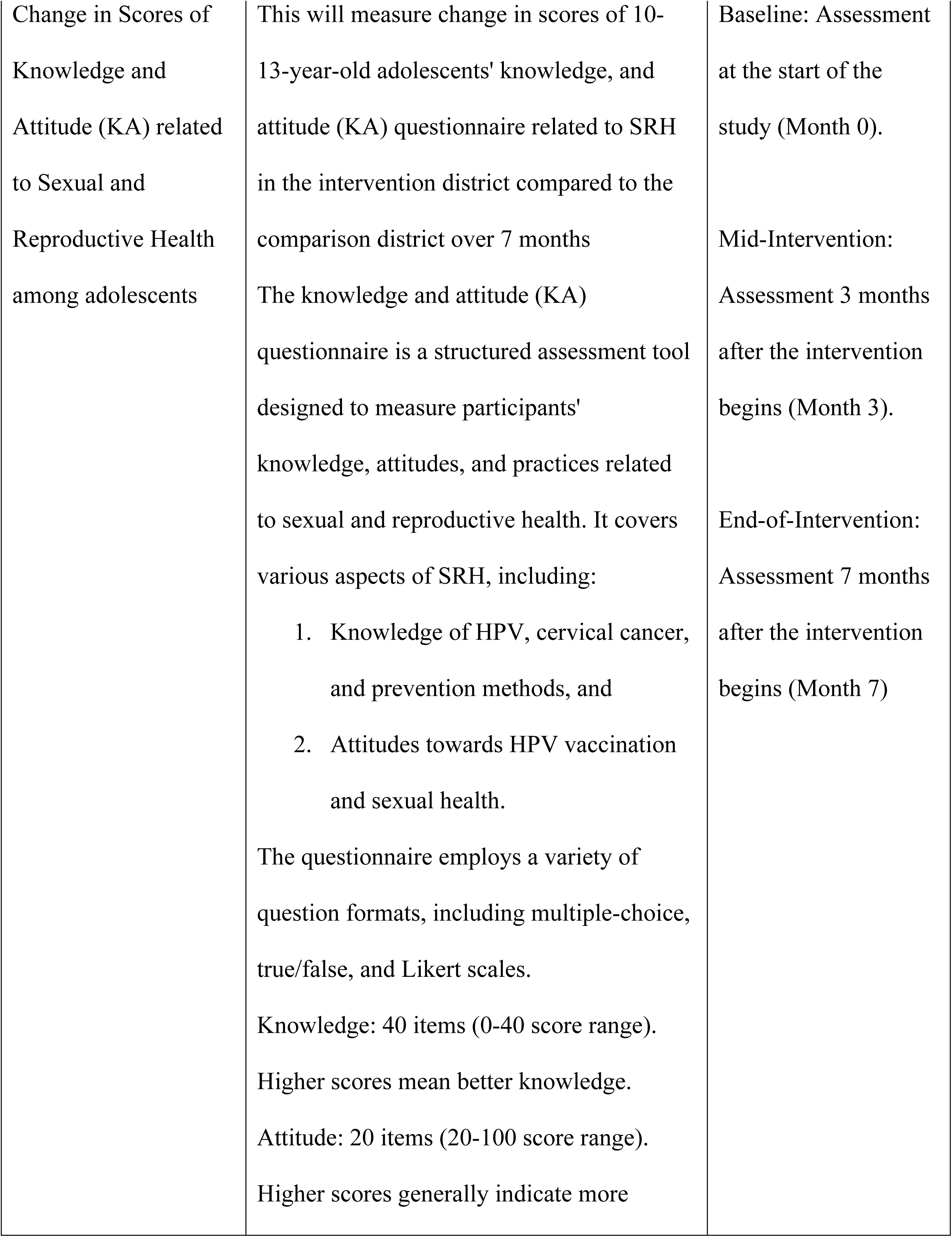
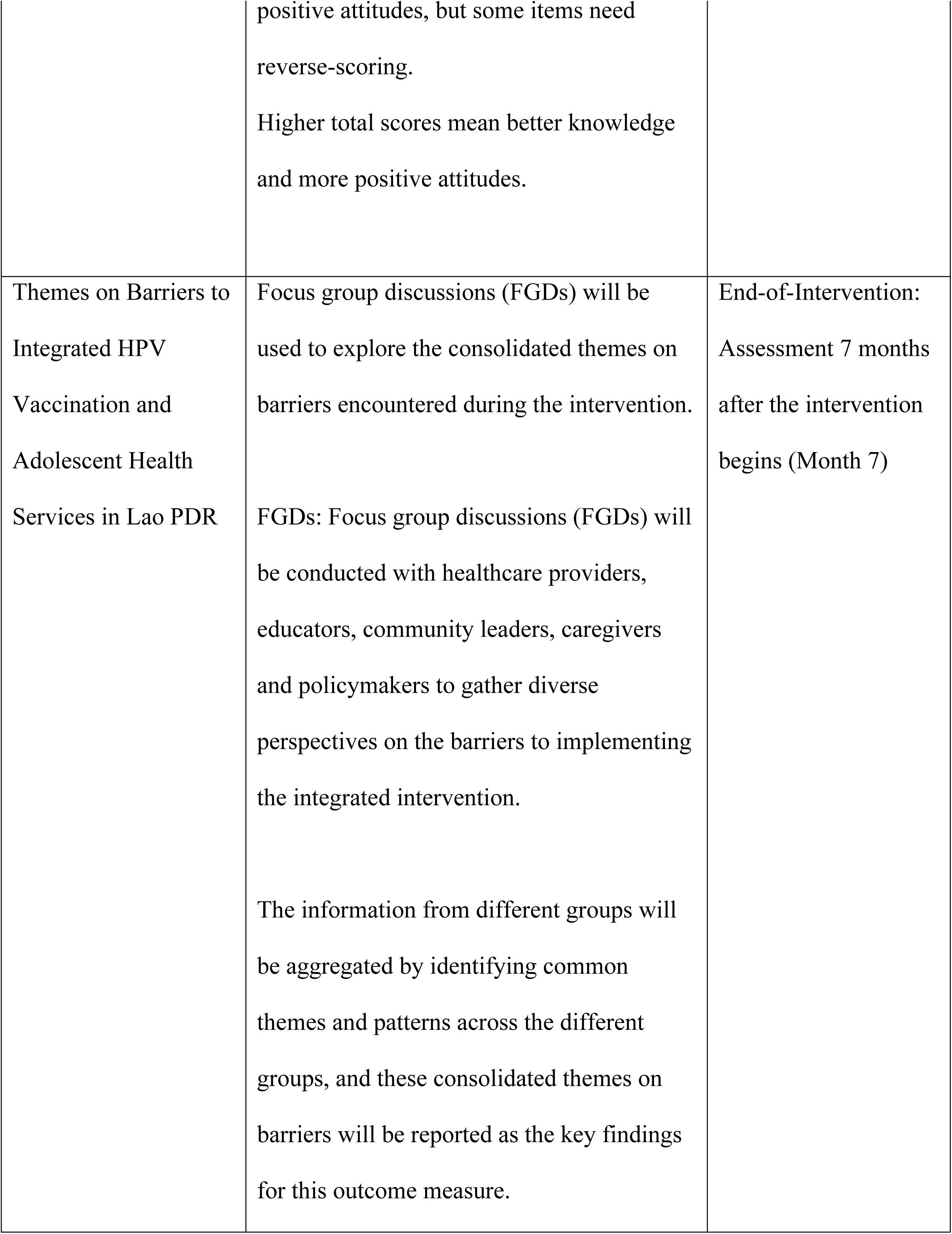
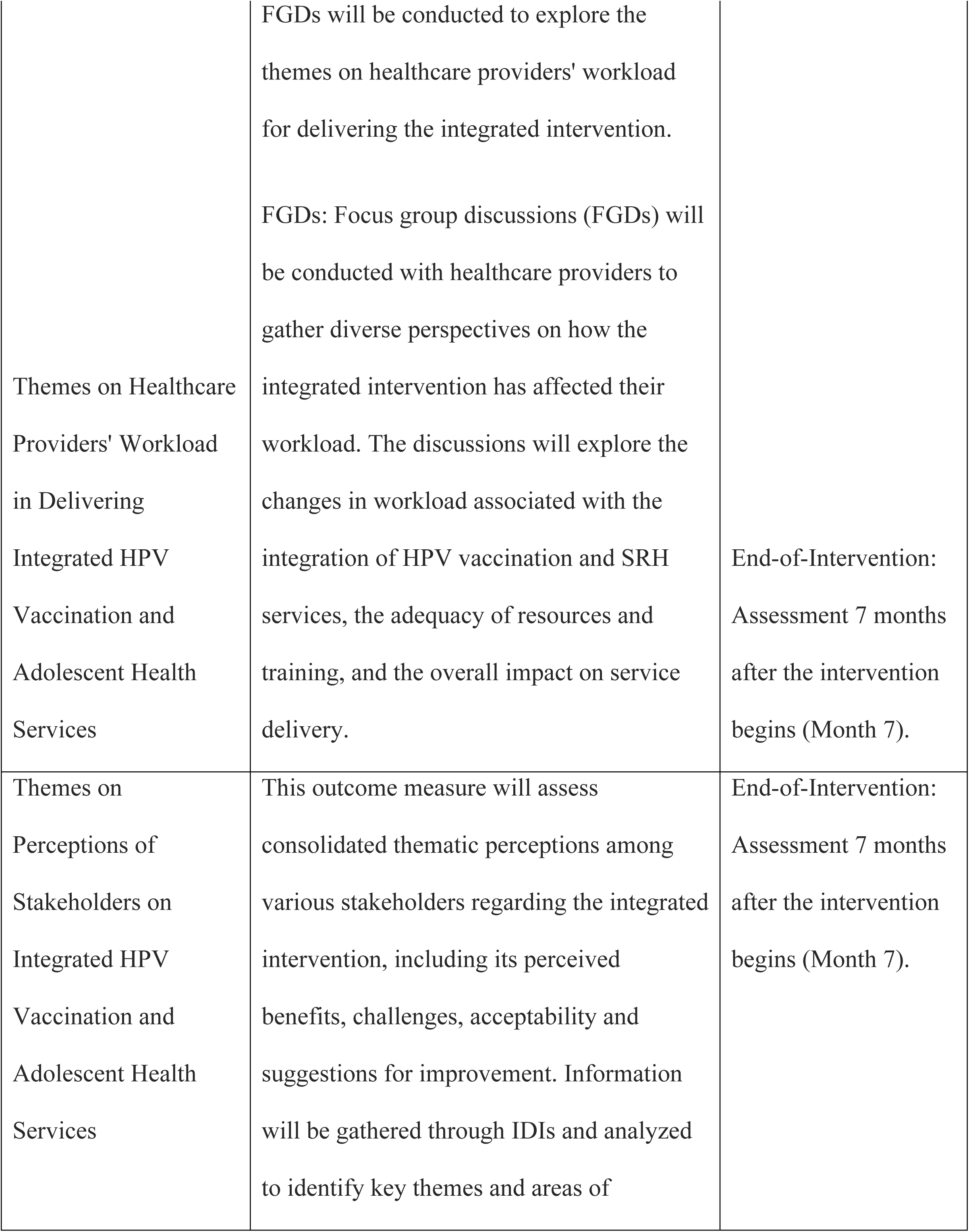
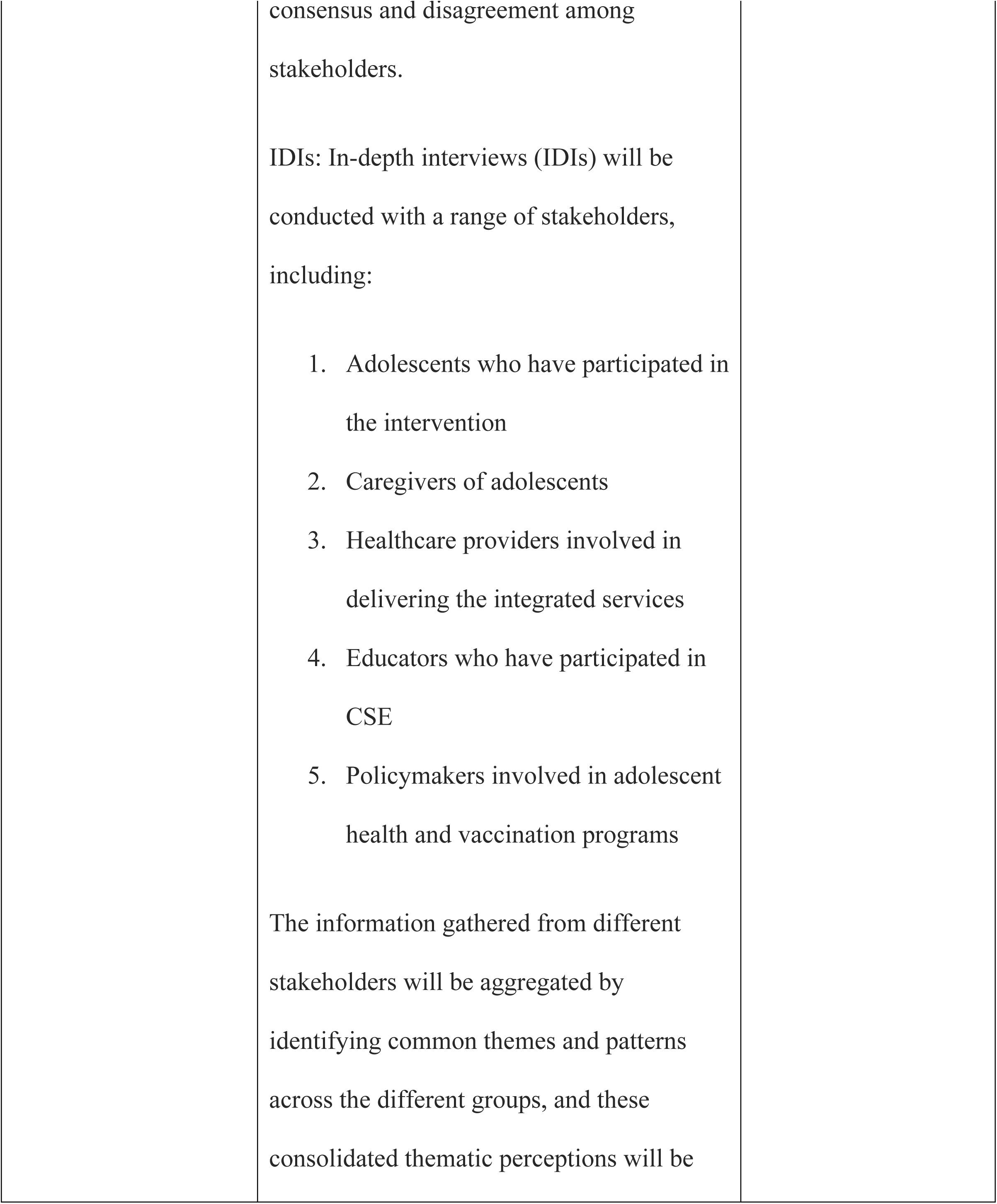
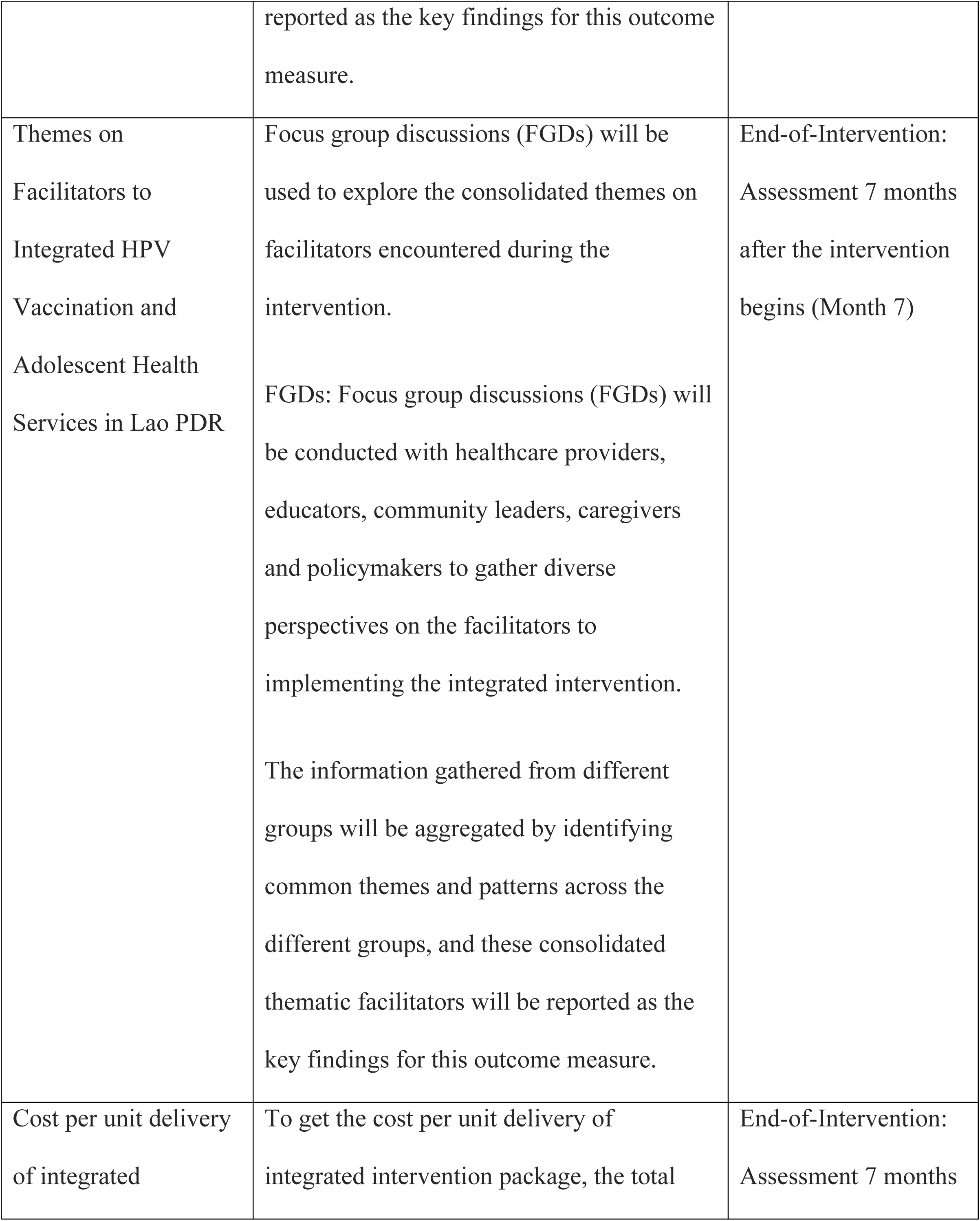
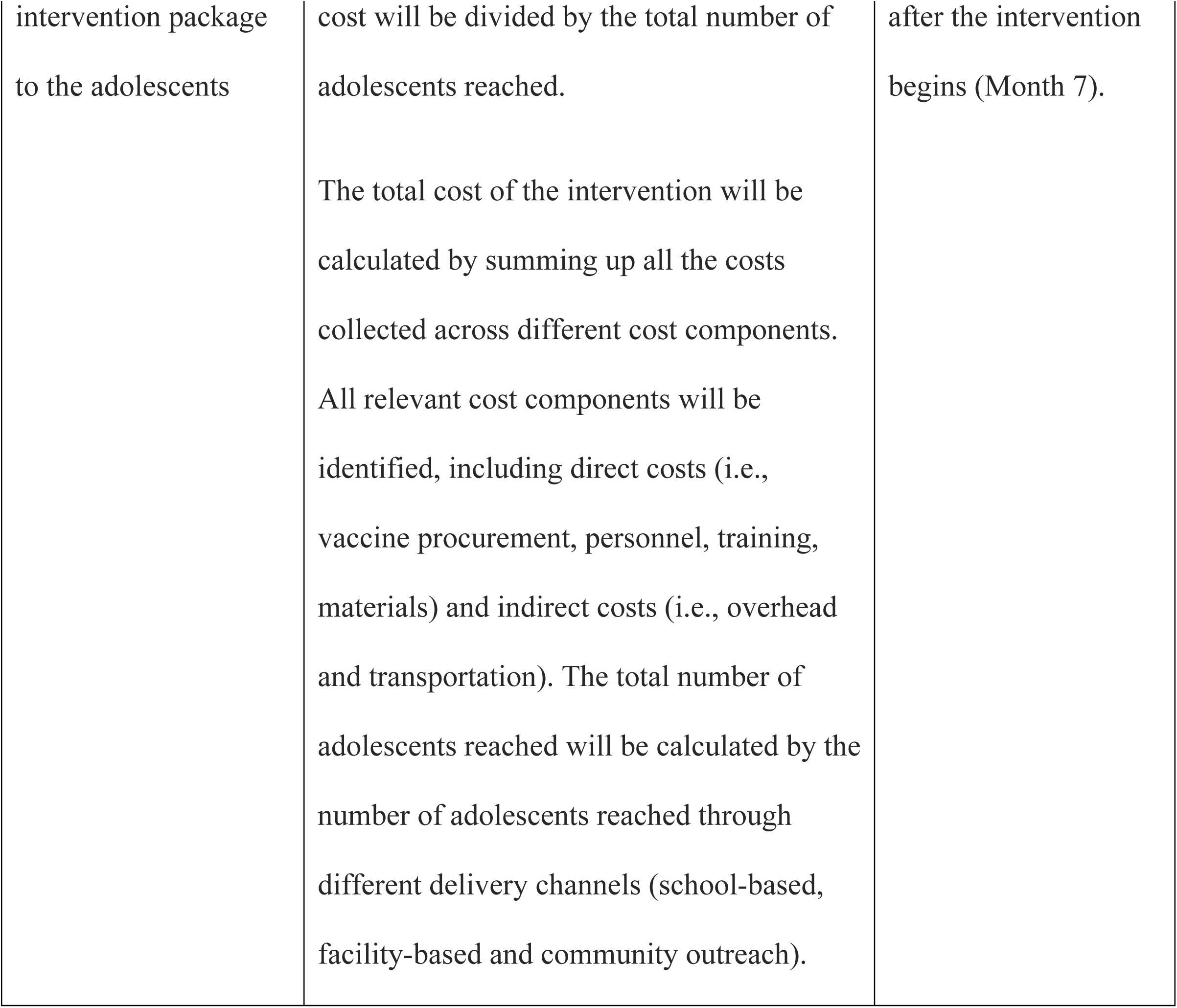
Outcome measures.

**Table 3.** Data quality control process.

|  |  |
| --- | --- |
| Service provider<br>(Primary Healthcare<br>staff, Educators) | ▪ Conduct integrated intervention with the target group at school and Health centres and record into the relevant forms |
| District Research<br>Facilitator | ▪ Collect the service record form and enter the data into digital form (Kobo) monthly<br>▪ Primary Conduct DQA to make sure the quality of data |
| Research Officer and M&E team | ▪ Track the submission weekly to ensure the integrated service data uploaded into system on time<br>▪ Track the progress of the programme according to the indicator to inform achievements and challenges. |
| Research Manager | ▪ Monthly DQA by dashboard and random checks<br>▪ Prepare the bi-annual project report and other reports (if any). Submit the report timely.<br>▪ Track the progress of the programme according to the indicator to inform achievements and challenges. |

To ensure impartiality, the outcomes of the study will be assessed by a team of investigators who are independent of the service providers, including health staff and schoolteachers. This independent assessment will provide an objective evaluation of the integrated intervention’s impact on HPV vaccine uptake and other adolescent health outcomes.

### 3.8 Data Analysis

To answer the research questions, a mixed-methods approach will be employed, utilizing both quantitative and qualitative data collection and analysis techniques.

#### 3.8.1 Quantitative Data

##### HPV Vaccine Uptake

To assess the impact of the integrated intervention on HPV vaccine uptake (research question 1.1), a difference-in-differences (DID) analysis will be employed to estimate the causal effect of the intervention, comparing the change in vaccination rates in the intervention district to the change in rates in the comparison district over the 7-month period. This approach helps to control for any pre-existing differences between the two districts and isolate the impact of the intervention. The McNemar Chi-Square statistic will be used for determining if there has been a significant change in vaccine uptake.

##### The plan for the *DID analysis*can be outlined as follows

###### Data Preparation

1. Outcome Variable: The primary outcome variable will be a binary indicator of whether a girl aged 10-13 has received the HPV vaccine (1 = vaccinated, 0 = not vaccinated). The data for this variable will be collected at baseline, 3^rd^ month, and endline in both the intervention and comparison districts.
2. Treatment Variable: The treatment variable will be a binary indicator of whether a girl resides in the intervention district (1 = intervention, 0 = comparison).
3. Time Variable: The time variable will be a binary indicator of whether the data was collected at endline (1 = endline, 0 = baseline or 3^rd^ month).
4. Covariates: The analysis will include relevant covariates that could potentially influence HPV vaccine uptake, such as age, socioeconomic status, education level, and access to healthcare. The selection of covariates will be based on existing literature and data availability.

###### DID Regression Model

The core of the DID analysis will involve estimating the following regression model:

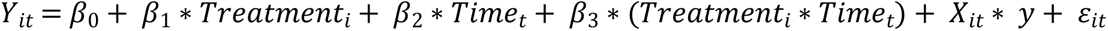

Where:

- *Y*_*it*_ = the outcome variable (HPV vaccination status) for individual i at time t.
- *Treatment*_*i*_ = the treatment variable indicating whether individual i is in the intervention or comparison district.
- *Time*_*t*_ = the time variable indicating whether the observation is at baseline, midline or endline.
- (*Treatment*_*i*_ ∗ *Time*_*t*_)= the interaction term between treatment and time, capturing the DID effect.
- *X*_*it*_ represents the vector of covariates for individual i at time t.
- *y* is the vector of coefficients for the covariates.
- *ɛ*_*it*_ is the error term.

###### Interpretation

The coefficient of primary interest is *β*_3_, which represents the DID estimate of the intervention’s impact on HPV vaccine uptake. A positive and statistically significant *β*_3_ would suggest that the integrated intervention led to an increase in vaccination rates in the intervention district compared to the comparison district.

###### Considerations for DID

○ Robustness Checks: The analysis will include robustness checks to assess the sensitivity of the results to different model specifications and assumptions. This may involve using alternative estimation methods, such as propensity score matching or synthetic control methods, to address potential confounding factors.
○ Subgroup Analysis: If the data allows, subgroup analyses will be conducted to explore whether the intervention’s impact varies across different groups of adolescents, such as those from different socioeconomic backgrounds or with varying levels of access to healthcare.
○ Qualitative Insights: The qualitative data collected in the study will be used to complement and enrich the quantitative findings, providing a deeper understanding of the mechanisms through which the integrated intervention influenced HPV vaccine uptake and other outcomes.

##### Knowledge and Attitudes (KA) Surveys

Statistic Repeated measures ANOVA will be used to evaluate the impact of the integrated intervention on adolescents’ knowledge, attitudes, and practices related to SRHR (research question 2.1), changes in KA scores between baseline and endline will be compared between the two districts to assess the impact of the intervention.

#### 3.8.2 Qualitative Data

The interviews and focus group discussions will be conducted in either the local language or English, based on the preference of the participants. The sessions will be audio-recorded to facilitate accurate transcription and ensure quality control. Transcripts originally in the local language will be translated into English for further analysis. The translated transcripts will then undergo coding using a deductive approach, employing NVivo software to manage and organize the data. The coded excerpts will be subsequently grouped into themes, reflecting the key patterns and insights emerging from the qualitative data. To enhance the richness and depth of the thematic analysis, direct quotes from the interview participants will be incorporated, ensuring anonymity to protect their privacy.

## 4 Ethical Considerations

### 4.1 Ethics Review

Ethical approval for the study has been received from the Research Ethics Committee (REC) in Laos with identification number of 833/REC dated 4th September 2024. Informed consent will be obtained from all participants or their legal guardians before any data collection. Confidentiality and privacy will be maintained throughout the study by anonymizing data and ensuring secure storage of information. The study will adhere to international ethical guidelines for research involving human subjects. Any potential risks or harms to participants will be minimized, and appropriate measures will be taken to ensure their well-being.

### 4.2 Informed consent

Informed consent for adolescent girls and boys participating in the integrated service implementation research in Lao PDR should consider their age, maturity, and understanding of the research. It is important to ensure that the adolescents are adequately informed and empowered to make decisions about their participation. The information provided to adolescent girls should be tailored to their age and level of understanding. Parental or guardian consent is required for to participate in research. Adolescents 10-13 years are needed to provide their own assent process, indicating their willingness to participate in the research. This process involves explaining the research to the adolescent in a way that they can understand and seeking their agreement to participate. In addition, the research team will ask the permission from schoolteachers as well. It is important to clarify the legal and ethical requirements for consent in Lao PDR and to involve parents or guardians as appropriate.

### 4.3 Risks and Benefits

There are not risks in participation in this study; however, participants may face stigma or discrimination related to HPV vaccination, particularly in communities where there is a lack of awareness or misconceptions about the vaccine.

The HPV vaccine has a well-established safety profile, with the most common side effects being local adverse events, primarily pain, redness, and swelling at the injection site, were commonly reported after HPV vaccination. Systemic reactions like headache and dizziness were also observed but were generally mild and self-limiting. Serious adverse events were rare, and no association was found between HPV vaccination and new-onset chronic diseases or infertility. (17)

There are no direct benefits in participating in this study, except small non-monetary such as refreshments, hygiene items etc and monetary compensation with no more than 5 USD per participant for their time in attending the surveys and interviews. Other potential benefits will be increased access to HPV vaccination, cost-effectiveness, and health system strengthening compelling arguments for integrated service implementation. The other indirect benefit is to increase the knowledge and attitude of SRHR among adolescent boys and girls. This research provides valuable insights for policymakers, healthcare practitioners, and researchers to optimize the integration of HPV vaccination services in Lao PDR and similar settings.

### 4.4 Confidentiality

Confidentiality is a critical aspect of participating in HPV vaccine integrated service implementation research in Lao PDR. There are several measures that should be in place to ensure confidentiality for participants such as data collected from participants should be anonymized to protect their identities. This may involve assigning unique identifiers to participants and using these identifiers in place of personal information. Any personal health information collected during the research should be stored securely, with access restricted to authorized personnel only.

### 4.5 Protocol amendments

Any significant changes to the study protocol will be communicated transparently to all relevant parties, including the ethics committee, funder and registries. Modifications to eligibility criteria, outcomes, or analyses are considered significant. The investigators will be informed directly, and the Research Ethics Committee (REC) in Laos will be notified to ensure the study remains ethically sound. Trial participants will be informed of any changes that might affect their participation or the study’s objectives. Updates will also be submitted to trial registries and journals to maintain accurate records and ensure transparency. The Principal

Investigator (PI) is required to notify the Secretary of the Research Ethics Committee (REC) in the event of any of the following:

- Significant changes to the project, including the reasons for the change and any ethical implications.
- Serious adverse effects on participants and the actions taken to address those effects.
- Unforeseen events or unexpected developments that require notification.
- Changes in research personnel, including the PI’s inability to continue in that role.
- The expiry of insurance coverage for the research project.
- Delays of more than 12 months in the commencement of the project.
- Termination or closure of the project.

## 5 Overall study timeline

The project workplan was finalized by Health Poverty Action (HPA), with the collaboration and agreement with implementation partners, University of Health Science (UHS) and National Immunization Program (NIP) through the series of consultation meeting. The work plan was aligned with research project management series, ensuring a) the feasibility under the local context and conditions of the country, b) relevance to influencing factors such as school calendars and community local calendars, c) availability resources including budget and d) willingness to actively participate by potential audiences. A simplified outline of research timeline can be seen in figure 3: timeline and content of intervention and comparison.

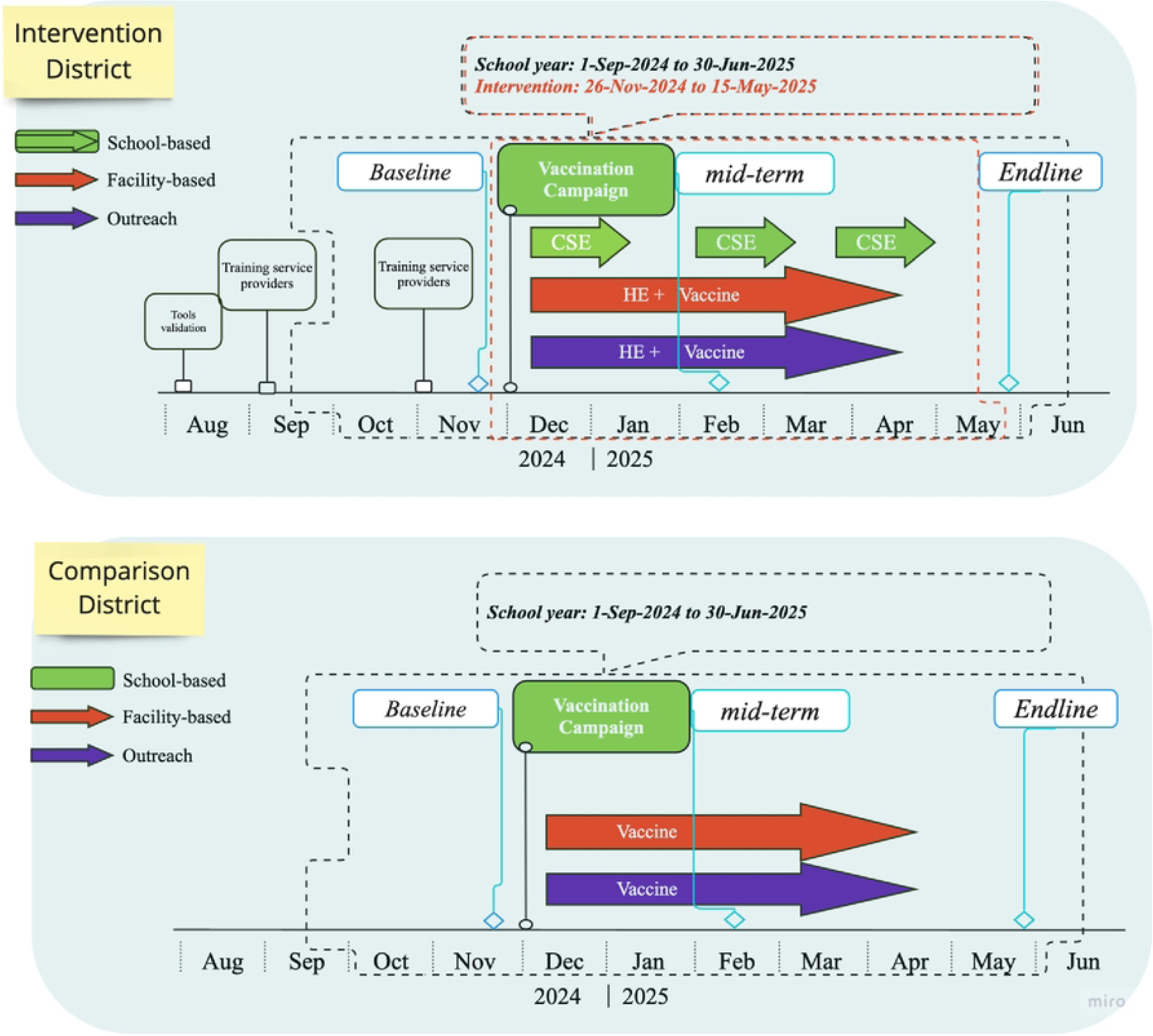

## 6 Discussion

### 6.1 Strengths and Limitation of the Study

This study has several strengths, including its mixed-methods design, which allows for a comprehensive understanding of the impact of integrating HPV vaccination with adolescent health services. The use of both quantitative and qualitative data will provide insights into the effectiveness of the intervention, as well as the barriers and facilitators to its implementation. Additionally, the study’s focus on adolescents, a vulnerable population with unique health needs, is a significant strength. By addressing the specific challenges faced by adolescents in accessing healthcare, this study has the potential to contribute to improved health outcomes for this group.

However, the study also has some limitations. The quasi-experimental design, while appropriate for this implementation setting, may not be as rigorous as a randomized controlled trial. This could limit the ability to draw definitive causal conclusions about the impact of the intervention. Additionally, the study’s focus on two districts in Lao PDR may limit the generalizability of the findings to other settings. Finally, the study’s reliance on self-reported data for some outcome measures could introduce bias.

### 6.2 Monitoring Plan

#### 6.2.1 Purpose of the plan

The Monitoring Plan is designed to ensure the effective collection, management, and analysis of data throughout the project. This plan aims to maintain high data quality, facilitate continuous learning, and support the achievement of project objectives. In the journey of integrating HPV vaccination with adolescent health services in Lao PDR, the Monitoring Plan will help track progress, identify challenges, and inform decision-making to improve health outcomes for adolescents.

#### 6.2.2 Timely production of quality data

Ensuring timely data production is a multifaceted challenge that extends beyond merely implementing electronic data recording forms and dashboards. One significant lesson from HPA’s operational research projects in the past is the lack of clear direction on who should use which forms and when. This often leads to confusion among field staff, resulting in inconsistent data collection practices. Additionally, the design of the forms themselves can be problematic. Poorly designed forms that are not user-friendly can hinder efficient data entry, causing delays and errors.

Another critical challenge is the lack of user confidence in using these electronic forms. Many field staff may not be familiar with digital data collection tools, leading to hesitancy and mistakes. This issue is compounded by a general lack of motivation among users, who may not see the immediate benefits of timely and accurate data entry. Without proper incentives and engagement, the enthusiasm for maintaining high data quality can wane.

Moreover, the timeliness and completion of data are sometimes not well-tracked. Optimal levels of motivation and technical aptitude within the central M&E team will be pivotal in mitigating such risk. Without a robust system to oversee the entire data management process, it becomes difficult to ensure that data is submitted on time and is complete. The central M&E team will be mobilized to provide the necessary technical skills to effectively use the interactive dashboard and other data management tools, further exacerbating the issue.

To address these challenges, the Monitoring Plan incorporates several solutions aimed at improving the overall data collection process:

- Clear Guidance and Training:

- Form Usage: A comprehensive guide will be developed to clearly outline who will use which forms, when, and how. This guide will be distributed to all field staff and stakeholders.
- Trainings: Regular, hands-on training sessions will be conducted to build user confidence in using electronic data recording forms. These sessions will cover form navigation, data entry, and troubleshooting common issues.
- Form Design:

- User-Friendly Design: Field staff will be consulted to design user-friendly forms that are intuitive and easy to navigate. The forms will be pilot tested, and feedback will be incorporated to improve usability.
- Standardization: Forms will be standardized to ensure consistency and ease of use across different data collection points.
- Motivation and Engagement:

- Regular Feedback: Feedback loops are to be established where field staff can share their experiences and challenges with the central M&E team. This will help in addressing issues promptly and keeping the team engaged.
- Tracking and Monitoring:

- Interactive Dashboard: An interactive data dashboard will be developed to provide real-time insights into data completion and timeliness. This dashboard will be accessible to all relevant stakeholders with monitoring responsibilities, enabling timely decision-making.
- Regular Check-ins: Regular check-ins will be conducted between the central M&E team and field staff to review data submission status and address any delays or issues.
- Capacity Building:

- Hands-on technical Training: Hands-on training will be provided by the Thematic Health Advisor and HPA Asia regional team for the central M&E team to enhance their technical aptitude in overseeing the data management system. This will include training on data validation, analysis, and dashboard management.
- Support System: A support system will be established where field staff and M&E team members can seek technical assistance and guidance as needed.

#### 6.2.3 Field Data Management and Quality Systems

HPA uses KOBO Toolbox to contribute to the Laotian national data management systems. Operational Research field staff will collect the HPV vaccine coverage and other relevant data from the primary health facilities and enter the data into KOBO toolbox in monthly. To ensure data accuracy, field staff will check primary data quality. Finally, M&E staff validate and store on KOBO server as well as analyse and support data information for report purposes.

To ensure the quality of data, HPA will pay attention to the following features of data and information:

- Validity and precision: by collecting the relevant information for the indicators, the HPA will clearly define the indicators, make sure that they are SMART indicators, define baseline and target and ensure the quality of the tools which are used for data collection.
- Reliability: to ensure the reliability of the data, all related M&E focal staff and field staff will be trained on using tools and data key-in and analysis.
- Integrity: HPA will create the linkage between different types of tools and different times of measurement if one indicator is measured several times.
- Procedure for validation: Data will be checked from different sources, verification means.
- Through regular trainings and multiple levels of checks, HPA ensures it maintains and improves its strong controls over collection, recording and reporting of high-quality data. The responsibility of each level in quality control is shown in below tables.

### 6.3 Dissemination Plan

The Dissemination Plan serves as a strategic roadmap for sharing our research findings on HPV integration with relevant stakeholders within Lao PDR and beyond. This plan outlines our objectives and identifies our target audience and methods of dissemination. It also provides an overview of the techniques we will use to disseminate our findings, the timeline for these activities, and how we plan to evaluate our efforts. As we are at the start of our research project, the methods and plan might still be adapted to better fit the context at different stages.

#### 6.3.1 Objectives

- To increase awareness about the importance and benefits of HPV vaccination and adolescent health services. We aim to ensure that the local community understands the research findings and their implications, and to encourage their active participation in the implementation of the vaccination program.
- To influence policy and practice by sharing our research findings with key stakeholders, including policymakers and health practitioners. We aim to provide evidence to support the integration of HPV vaccination with adolescent health services, demonstrating its potential to increase HPV vaccine coverage and contribute to adolescent health outcomes.
- To contribute to the international body of knowledge on HPV vaccination and adolescent health services. We aim to share our findings with the global research community and international health organizations, providing evidence that can be used to inform similar initiatives in other countries.

#### 6.3.2 Target Audiences

The key target audiences for this dissemination plan are:

- Local community members, including young adolescents, their parents and direct community members, as well as related community health workers, district and provincial staff to increase awareness about the importance and benefits of HPV vaccination and adolescent health services.
- National stakeholders, such as policymakers and health practitioners from the Ministry of Health and the Ministry of Education, influence policy and practice.
- Global research community and international health organizations, including GAVI and WHO to contribute to the international body of knowledge on HPV vaccination and adolescent health services.

#### 6.3.3 Dissemination Methods

- IEC/BCC Materials: Information, Education, and Communication (IEC) and Behaviour Change Communication (BCC) materials will be used to share key messages and findings. These could include brochures, posters, and leaflets that can easily be distributed and understood by the community.
- Training workshops: Throughout the programme delivery, we will deliver training workshops for health practitioners and community health workers on the integration of services co-delivered with HPV vaccination.
- Policy Brief: A brief will be prepared to be shared with policymakers in Lao PDR. This will summarize our findings and provide recommendations for policy for advantages of HPV integration with other health services and/or through different delivery methods.
- Workshops: We will organize workshops for key stakeholders, including those from the relevant ministries and departments. This will provide an opportunity to share our research, discuss the findings, and facilitate knowledge and learning exchange among skey stakeholders.
- Academic Publication: We will aim to publish our research findings in academic journals. This will ensure they reach the wider research community and contribute to the global body of knowledge on HPV vaccination integration with SRHR education.
- Narrative Reports: We will prepare narrative reports that detail our research process, findings, and implications. These can be shared with a wide range of stakeholders.
- International Conferences: We will present our findings at international conferences (and GAVI Global meetings). This will provide an opportunity to share our research with a global audience, receive feedback, and network with other researchers in the field.

## 7 Role of study funders

Funder of the study, Gavi, provides an overarching aim of the learning agenda. Gavi, the Vaccine Alliance, commissions a set of implementation research projects to contribute evidence on feasible, effective, integrated service approaches for adolescents to increase and sustain HPV vaccination access, equity, and coverage in priority countries. to test how integrated approaches could increase HPV coverage equitably and sustainably, as well as contribute to additional, applicable adolescent health outcomes.

The conceptualization of the study in Laos PDR, design of its study methodology, intervention, study procedures and analysis are decided by the authors. Gavi makes comments on the study protocol, methodology and intervention design. Authors make the decision to submit the protocol and findings for publication.

## 8 Authors’ contributions

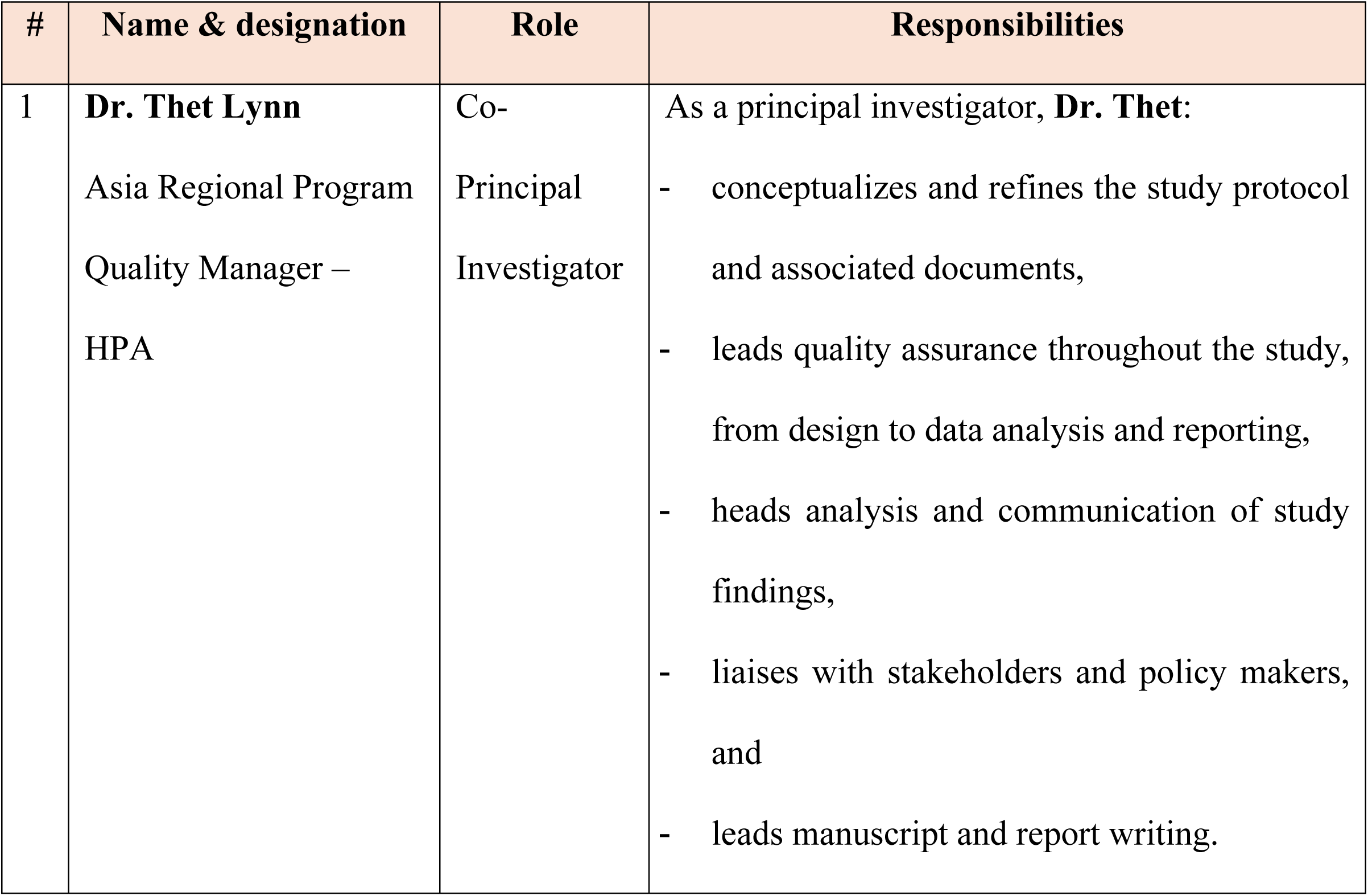
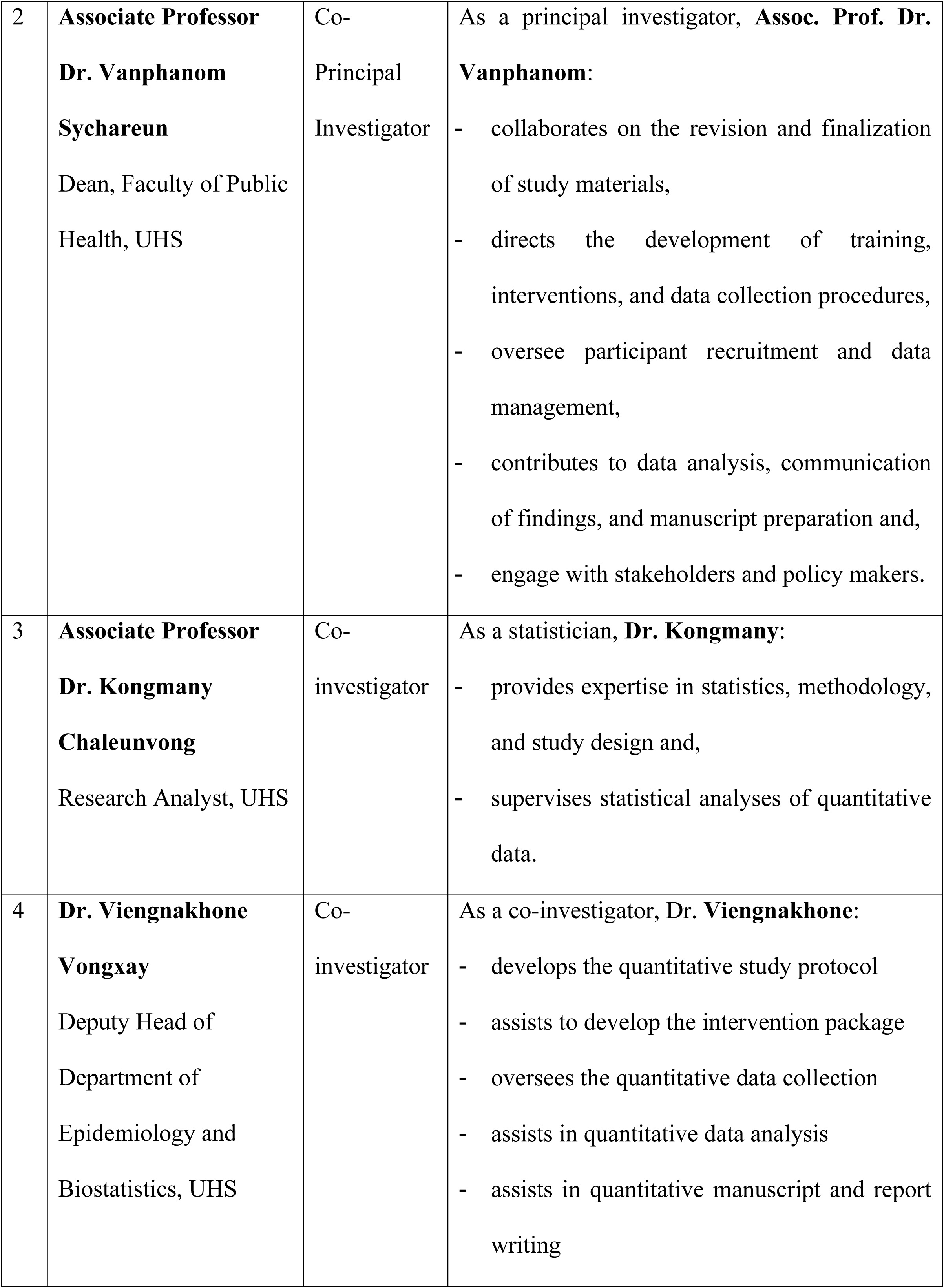
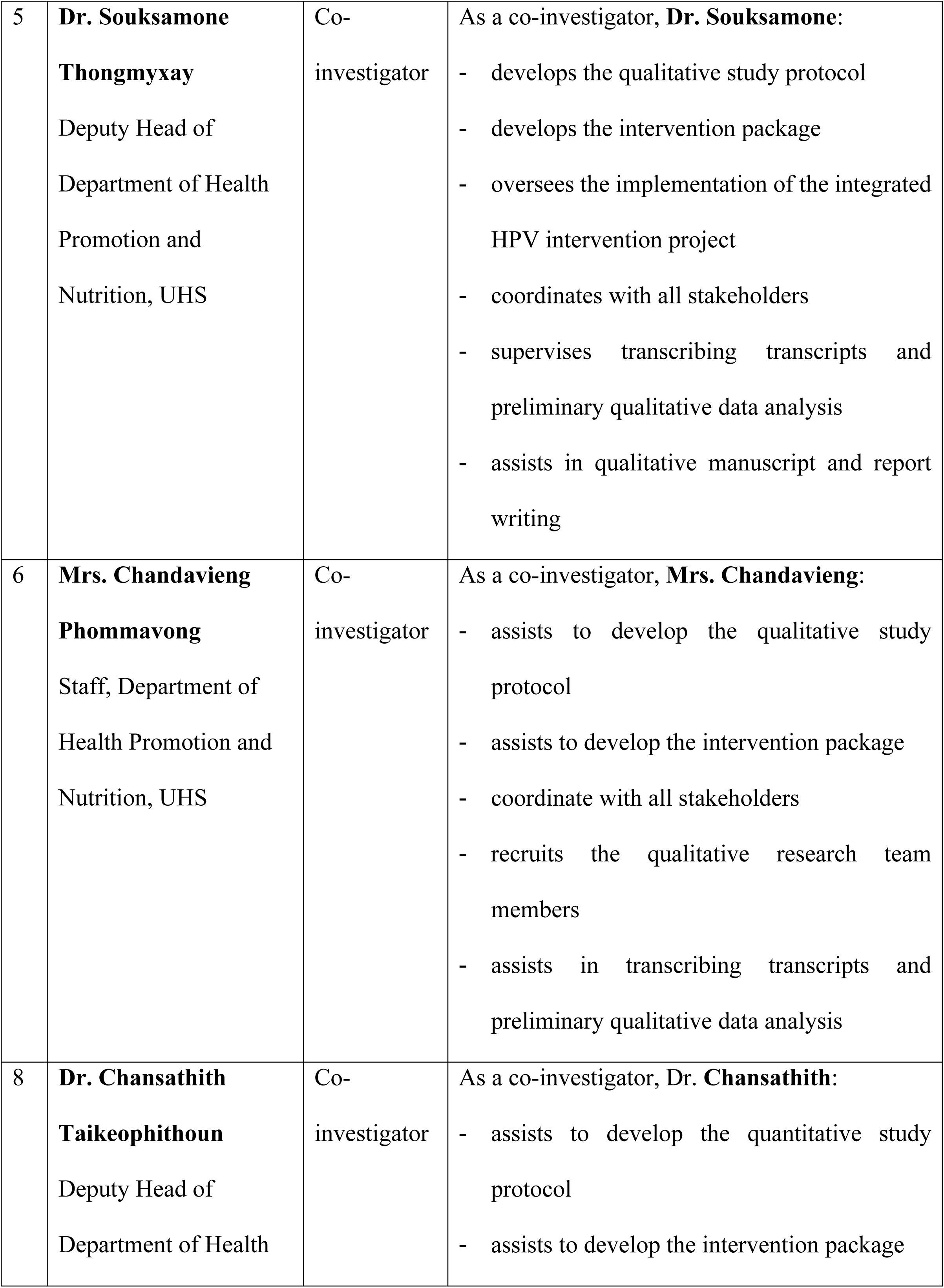
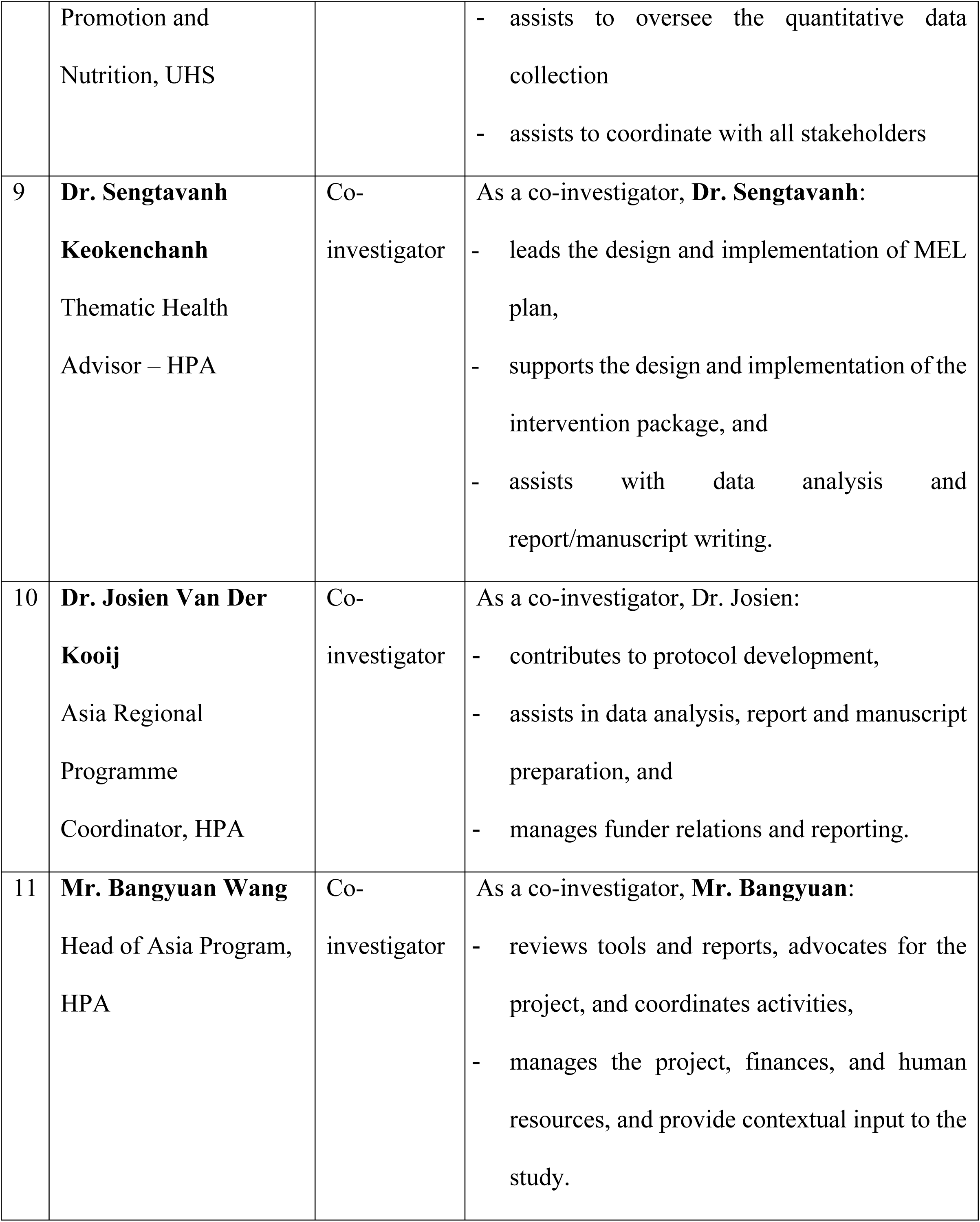

## Data Availability

No datasets were generated or analysed during the current study. All relevant data from this study will be made available upon study completion.

